# Plasma proteomics reveals stage-dependent biological remodelling in Alzheimer’s disease

**DOI:** 10.64898/2026.09.14.26362968

**Authors:** Yuna Gu, Jun Pyo Kim, Bo Hyun Kim, Hyunwoo Lee, Jungah Lee, Heekyoung Kang, Sohyun Yim, Daeun Shin, Hee Kyung Park, Seonghyeon Kim, Byung Hwa Lee, Jeongha Lee, Hyeongjin Kim, Henrik Zetterberg, Min Young Chun, Duk L. Na, Jae-Hong Lee, Soo Hyun Cho, Byeong C. Kim, Si Eun Kim, Gyeongmo Sohn, Jihwan Yun, Jae-Sung Lim, Juyoung Kim, Kyungbok Lee, Jaeho Kim, Hee Jin Kim, Eun-Joo Kim, Hyemin Jang, Sang Won Seo

## Abstract

Plasma biomarkers of amyloid-associated tau phosphorylation (T1) and established tau proteinopathy (T2) can approximate Alzheimer’s disease stage, but whether plasma-defined stages correspond to broader biological states is unknown. In 1,035 participants from a multicentre Korean cohort, we used a 220-plex immunoassay platform to compare T1/T2 biomarkers against amyloid and tau positron emission tomography anchors and construct a five-stage framework. Brain-derived phosphorylated tau 217 and endogenously cleaved microtubule-binding-region tau formed the parsimonious backbone. Baseline stage ordered Clinical Dementia Rating–Sum of Boxes trajectories, and forward within-person stage movement was associated with faster clinical worsening. Among 197 non-tau proteins, 34 were stage-associated; the T1-positive transition showed the broadest proteomic shift, with more selective remodelling later. In serial samples, 15 proteins changed longitudinally, with 13 recapitulating the cross-sectional stage pattern. Thus, plasma-defined disease position mapped onto distinct, partly dynamic biological states beyond the defining T1/T2 biomarkers.

---

Alzheimer’s disease (AD) biological staging aims to locate individuals along the continuum of AD neuropathologic change. The revised criteria distinguish early-changing Core 1 biomarkers from later-changing Core 2 biomarkers, while positron emission tomography (PET) provides an established reference for increasing amyloid and tau burden.^1–4^ Blood-based assays offer a more scalable route to estimate disease position.^5,6^ Recent studies have established the chronology of plasma T1 and T2 analytes, a sequential phosphorylated tau 217 (p-tau217)/ endogenously cleaved microtubule-binding-region tau (eMTBR-tau) diagnostic workflow and plasma approximation of PET-based stages.^7–11^ Whether such plasma-defined disease positions correspond to broader biological states beyond the defining biomarkers remains unknown.

Within this architecture, T1 biomarkers are phosphorylated and secreted AD tau fragments that become abnormal around amyloid PET positivity, whereas T2 biomarkers change later and more closely reflect established AD tau proteinopathy.^1,7,8^ Candidate T1 measures include p-tau181, p-tau212, p-tau217 and p-tau231, with conventional and brain-derived (BD) formats available for several phosphosites; candidate T2 measures include p-tau205, BD-p-tau205 and eMTBR-tau.^7,12–14^ Several single-marker and multianalyte representations could therefore be used to construct a plasma staging backbone. A common high-plex platform enables these candidates to be compared under the same analytical conditions.

Biological staging and broader biological characterisation are complementary but distinct tasks. Complementary T1 and T2 plasma biomarkers define disease position, whereas markers of neuronal injury, inflammation, synaptic dysfunction and other pathophysiological processes describe the biological state accompanying that position.^1,15–21^ A plasma staging hierarchy would gain biological support if broader proteomic profiles varied systematically across its stages, if differences were localised to particular adjacent-stage boundaries and if part of the cross-sectional pattern was recapitulated within individuals over time. High-plex profiling makes it possible to test these features independently of marker selection.

In the multicentre K-ROAD cohort, we used a 220-plex immunoassay platform (NULISAseq) to build, validate and characterise a plasma biological staging framework. We first compared prespecified T1 and T2 candidates and multianalyte representations against amyloid and nested tau PET anchors to build a parsimonious five-stage backbone. We then validated the hierarchy using longitudinal CDR-SB trajectories, all four adjacent-stage slope contrasts and within-person stage movement. Finally, we characterised stage-associated biology across 197 non-tau proteins, localised differences across adjacent stages and tested longitudinal recapitulation in serial plasma samples.

## Results

### Cohort and baseline characteristics

The baseline staging cohort comprised 1,035 participants (mean age 71.5 years [SD 8.2]; 654 [63.2%] female), including 268 (25.9%) cognitively unimpaired participants, 504 (48.7%) with mild cognitive impairment and 263 (25.4%) with dementia; 692 (66.9%) were Aβ PET-positive (Table 1). Of the 1,035 participants, 1,007 (97.3%) were stageable and 28 (2.7%) had T1-negative/T2-positive profiles and were therefore unstageable. Across Stages 0–4, median CDR-SB increased from 0.5 to 4.5, while Aβ PET positivity increased from 10.3% to 95.0%, with the largest shift in Aβ positivity occurring between No plasma tau abnormality and Tau phosphorylation (Table 1).

**Table 1.** Baseline demographic, clinical, and Aβ PET characteristics by plasma biological stage.

| Characteristic | Overall<br>(n=1,035) | Stage 0<br>No plasma tau abnormality<br>(n=311) | Stage 1<br>Tau phosphorylation<br>(n=206) | Stage 2<br>Early aggregation<br>(n=99) | Stage 3<br>Moderate aggregation<br>(n=191) | Stage 4<br>Advanced aggregation<br>(n=200) | Unstageable<br>T1-/T2+<br>(n=28) |
| --- | --- | --- | --- | --- | --- | --- | --- |
| Age, years | 71.5 (8.2) | 70.3 (7.9) | 74.3 (6.9) | 75.9 (6.4) | 72.9 (6.7) | 66.4 (9.4) | 74.0 (6.3) |
| Female sex, n (%) | 654 (63.2) | 185 (59.5) | 119 (57.8) | 68 (68.7) | 121 (63.4) | 142 (71.0) | 19 (67.9) |
| Education, years | 11.8 (4.5) | 11.8 (4.4) | 12.2 (4.8) | 10.5 (4.5) | 11.8 (4.4) | 12.1 (4.2) | 11.0 (4.5) |
| APOE ε4 carrier, n (%) | 488 (47.2) | 75 (24.2) | 113 (54.9) | 64 (64.6) | 126 (66.3) | 102 (51.0) | 8 (28.6) |
| **Clinical diagnosis, n (%)** |  |  |  |  |  |  |  |
| Cognitively unimpaired | 268 (25.9) | 194 (62.4) | 43 (20.9) | 9 (9.1) | 10 (5.2) | 1 (0.5) | 11 (39.3) |
| Mild cognitive impairment | 504 (48.7) | 103 (33.1) | 136 (66.0) | 66 (66.7) | 108 (56.5) | 79 (39.5) | 12 (42.9) |
| Dementia | 263 (25.4) | 14 (4.5) | 27 (13.1) | 24 (24.2) | 73 (38.2) | 120 (60.0) | 5 (17.9) |
| MMSE score | 26.0 [22.0–28.0] | 28.0 [26.0–29.0] | 26.5 [24.0–28.0] | 24.0 [21.0–27.0] | 23.0 [21.0–26.0] | 20.0 [16.0–23.0] | 27.0 [25.0–29.0] |
| CDR-SB | 2.0 [0.5–3.5] | 0.5 [0.5–1.0] | 1.5 [0.5–2.5] | 3.0 [1.0–4.0] | 3.0 [1.5–4.5] | 4.5 [3.0–7.0] | 1.0 [0.5–2.0] |
| Aβ PET Centiloid | 58.6 [8.7–84.8] | 3.4 [–3.2–10.6] | 64.6 [44.0–81.4] | 70.2 [53.1–88.5] | 79.1 [63.6–94.4] | 82.5 [66.5–104.8] | 6.6 [1.6–30.3] |
| Aβ PET positive, n (%) | 692 (66.9) | 32 (10.3) | 185 (89.8) | 93 (93.9) | 183 (95.8) | 190 (95.0) | 9 (32.1) |

Data are mean (SD), median [IQR], or n (%). Overall includes all 1,035 participants; stage-specific analyses used the 1,007 stageable participants. The unstageable group comprised T1-negative/T2-positive profiles. Aβ PET positivity was defined as Centiloid >24. MMSE was available in 1,022 participants and APOE genotype in 1,033.
APOE $\epsilon$ 4 carrier denotes E2/E4, E3/E4, or E4/E4 genotype.
Abbreviations: Aβ , amyloid-β ; APOE, apolipoprotein E; CDR-SB, Clinical Dementia Rating–Sum of Boxes; IQR, interquartile range; MMSE, Mini-Mental State Examination; PET, positron emission tomography; T1, amyloid-associated phosphorylated and secreted AD tau; T2, established AD tau proteinopathy.

### Building a parsimonious plasma staging backbone

Among T1 candidates, BD-p-tau217 showed the highest discrimination for Aβ PET positivity, with an area under the receiver-operating-characteristic curve (AUC) of 0.936, exceeding conventional p-tau217 (AUC 0.925; ΔAUC 0.011, 95% CI 0.004–0.018); a four-marker BD composite performed less well (AUC 0.929; Fig. 2a, Supplementary Fig. S1 and Supplementary Table S1). For T2, eMTBR-tau outperformed p-tau205 and BD-p-tau205 across the Temporal Meta, neocortical composite (NeoC) and high-NeoC tau PET anchors (AUCs 0.986, 0.975 and 0.969, respectively; Fig. 2b and Supplementary Table S2). Adding BD-p-tau205 provided no consistent improvement, including a ΔAUC of −0.040 for the high-NeoC anchor (Supplementary Fig. S2a and Supplementary Table S2). eMTBR-tau increased across the nested PET-defined hierarchy, with the frozen thresholds separating tau PET-negative, Early aggregation, Moderate aggregation and Advanced aggregation groups (Supplementary Fig. S2b).

**Figure 1.**
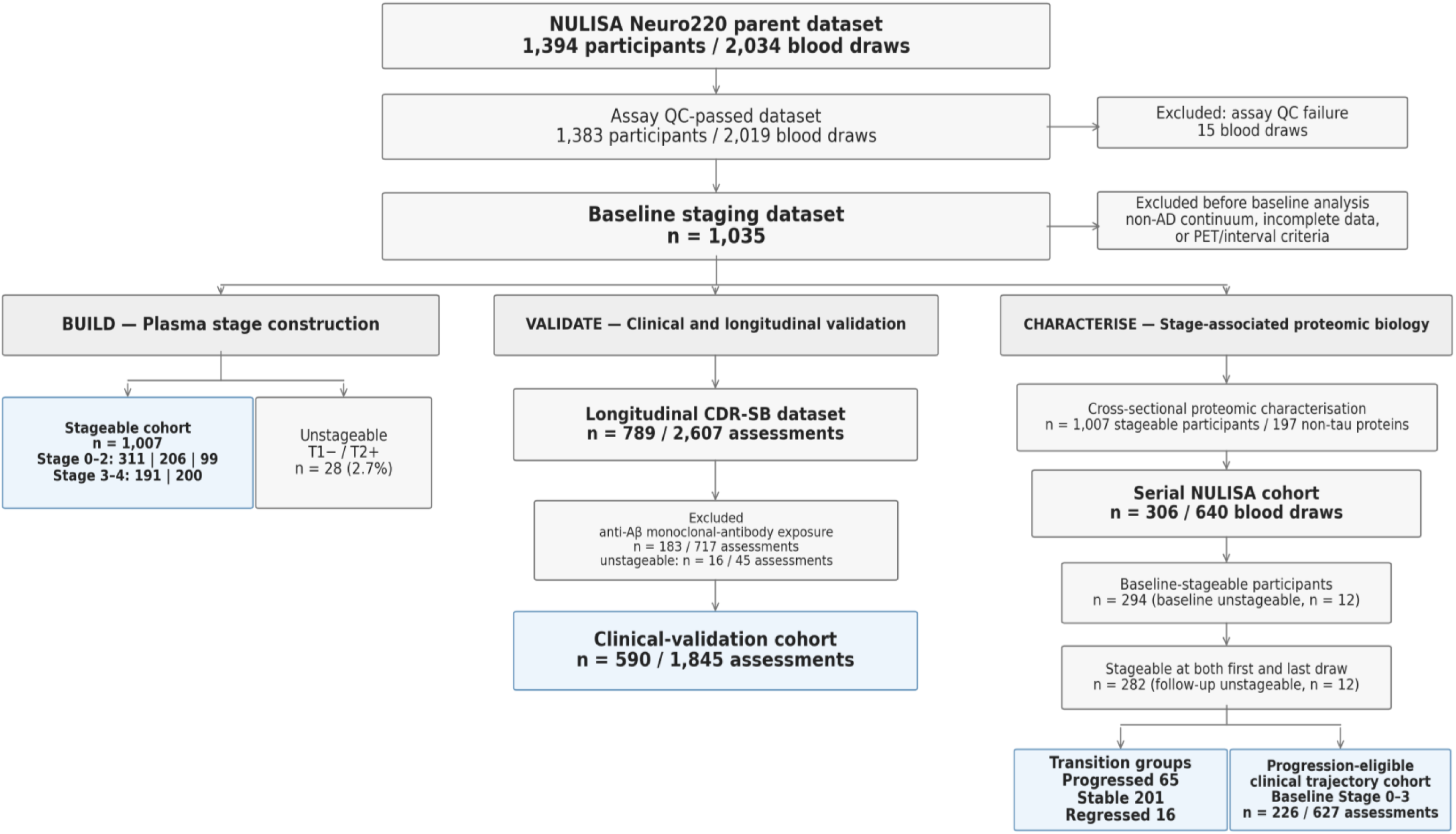
Build, validate and characterise: cohort flow and analytic datasets. Overlapping datasets were used to construct the frozen T1/T2 plasma stages, validate baseline clinical trajectories and within-person stage movement, and characterise cross-sectional and serial non-tau proteomic patterns. Numbers and exclusions are shown in the diagram; the progression-eligible clinical trajectory cohort additionally required baseline Stage 0–3 and eligible longitudinal CDR-SB assessments. CDR-SB, Clinical Dementia Rating–Sum of Boxes; NULISA, nucleic acid-linked immuno-sandwich assay; T1, amyloid-associated phosphorylated and secreted AD tau; T2, established AD tau proteinopathy.

**Figure 2.**
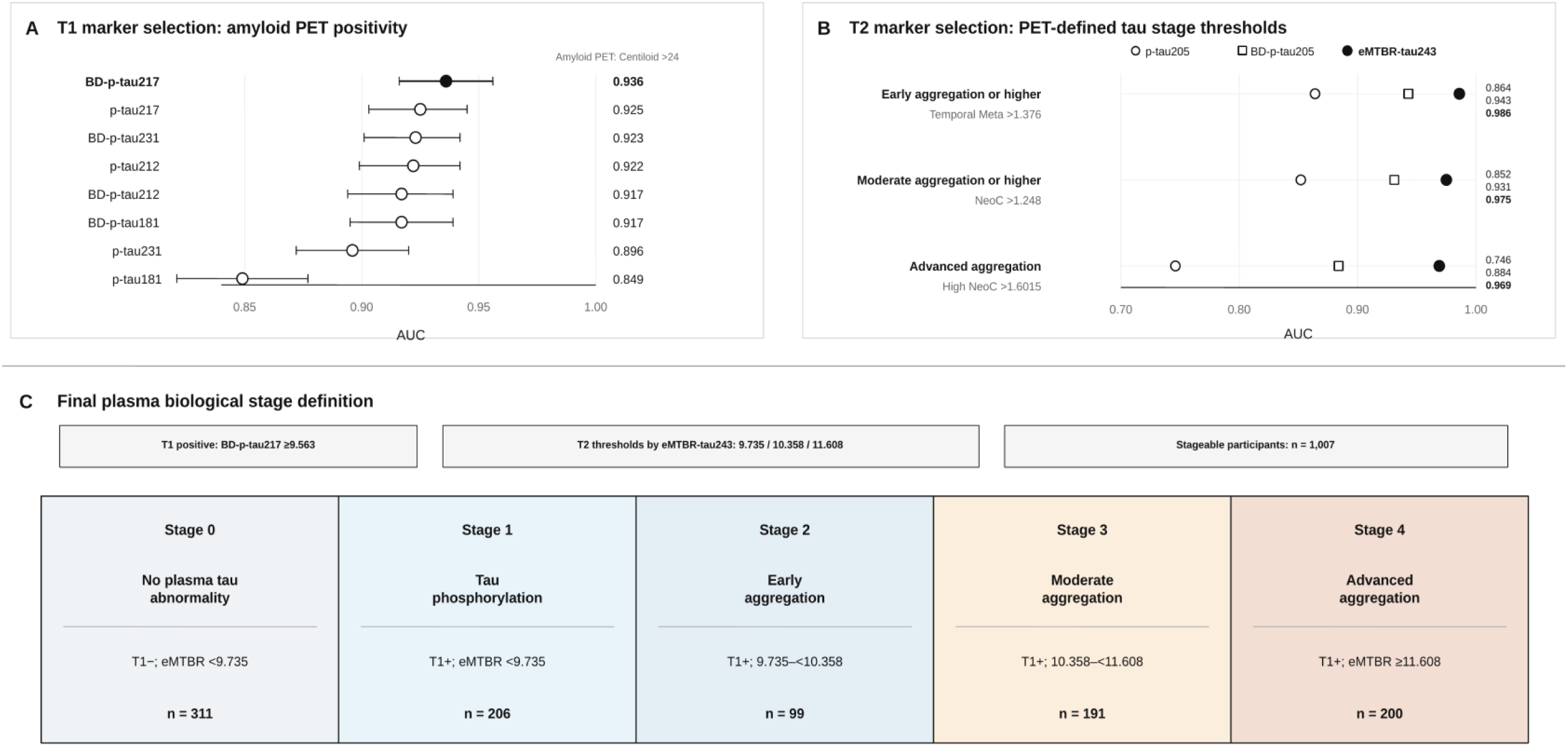
PET-anchored construction of the plasma biological staging framework. (a) T1 candidates were benchmarked against Aβ PET positivity (Centiloid >24); BD-p-tau217 showed the highest AUC and was selected at 9.563 log2 NPQ. (b) T2 candidates were benchmarked against Temporal Meta >1.376, NeoC >1.248 and high NeoC >1.6015; eMTBR-tau showed the highest AUCs and yielded thresholds of 9.735, 10.358 and 11.608. (c) Frozen Stage 0–4 hierarchy and unstageable T1-negative/T2-positive profile. Cognition and non-tau proteins were excluded from construction. Aβ, amyloid-β; AUC, area under the receiver-operating-characteristic curve; BD, brain-derived; eMTBR-tau, endogenously cleaved microtubule-binding-region tau243; NeoC, neocortical composite; NPQ, NULISA Protein Quantification; PET, positron emission tomography; T1, amyloid-associated phosphorylated and secreted AD tau; T2, established AD tau proteinopathy.

The final algorithm used BD-p-tau217 ≥9.563 for T1 positivity. Stage 0 (No plasma tau abnormality) was T1-negative with eMTBR-tau <9.735; T1-positive participants were classified as Stage 1 (Tau phosphorylation; eMTBR-tau <9.735), Stage 2 (Early aggregation; eMTBR-tau 9.735–<10.358), Stage 3 (Moderate aggregation; eMTBR-tau 10.358–<11.608) or Stage 4 (Advanced aggregation; eMTBR-tau ≥11.608). Counts were 311, 206, 99, 191 and 200, respectively; 28 participants with T1-negative/eMTBR-tau ≥9.735 profiles were unstageable (Fig. 2c).

### Clinical and longitudinal validation

Ordinal stage correlated with baseline CDR-SB (Spearman ρ=0.649; P<0.001). In 590 participants with 1,845 assessments, estimated annual CDR-SB slopes were 0.191, 0.382, 0.674, 1.177 and 1.943 points per year across No plasma tau abnormality, Tau phosphorylation, Early aggregation, Moderate aggregation and Advanced aggregation, respectively (stage-by-time χ²(4)=271.386, P=1.60×10⁻⁵⁷). All four prespecified adjacent-stage slope contrasts remained significant after false-discovery-rate correction, with q=0.0466, 0.0330, 4.73×10⁻⁴ and 9.63×10⁻¹⁰, respectively (Fig. 3a and Supplementary Table S3).

**Figure 3.**
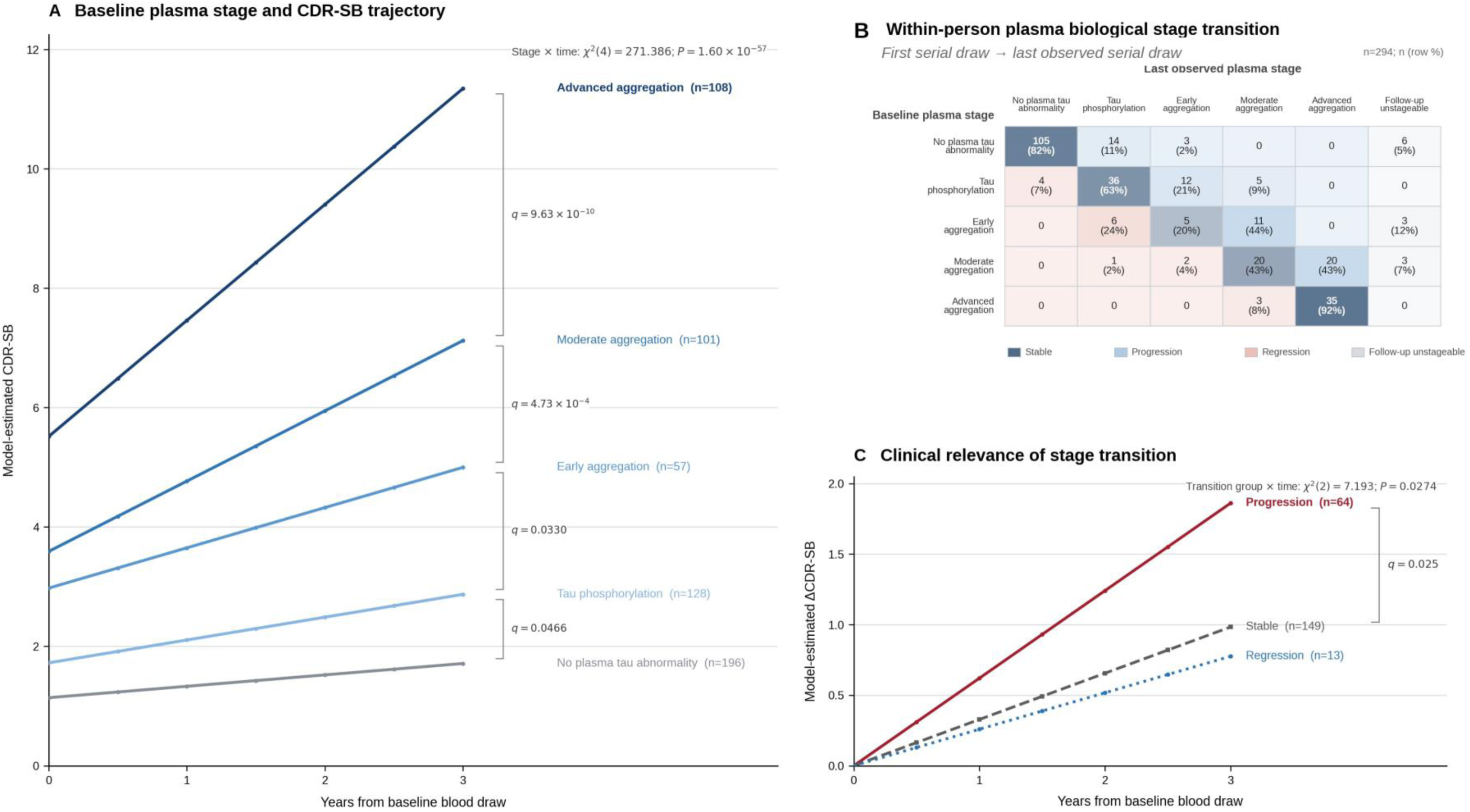
Clinical and longitudinal validation. (a) Model-estimated CDR-SB trajectories in 590 participants with 1,845 assessments; brackets show the four BH-FDR-adjusted adjacent-stage slope contrasts. (b) First-to-last stage matrix in 294 baseline-stageable participants; 12 were unstageable at follow-up. Of 282 participants stageable at both draws, 65 progressed, 201 remained stable and 16 regressed; 80.2% of directional changes were forward. (c) Trajectories by transition group in 226 participants with 627 assessments; progression versus stability, +0.293 points per year, q=0.025. Panel c reflects longitudinal coupling rather than landmark prediction. BH-FDR, Benjamini– Hochberg false discovery rate; CDR-SB, Clinical Dementia Rating–Sum of Boxes.

Among 282 participants stageable at both serial draws, 65 progressed, 201 remained stable and 16 regressed; 65 of 81 directional changes were forward (80.2%; P=3.64×10⁻⁸; Fig. 3b). In the progression-eligible cohort (n=226; 627 assessments), progressors worsened faster than stable participants (+0.293 CDR-SB points per year, 95% CI 0.062–0.523; q=0.025), whereas regression did not differ from stability (Fig. 3c).

### Biological characterisation of plasma-defined stages

Among 197 quality-control (QC)-passed non-tau proteins, 34 were associated with categorical stage after age and sex adjustment and Benjamini–Hochberg false-discovery-rate (BH-FDR) correction (Fig. 4a and Supplementary Table S4). The strongest associations included glial fibrillary acidic protein (GFAP), acetylcholinesterase (ACHE), neurofilament light chain (NfL), growth-associated protein 43 (GAP43), β-site amyloid precursor protein cleaving enzyme 1 (BACE1) and secreted phosphoprotein 1 (SPP1). Adjacent-stage contrasts localised the broadest differences between No plasma tau abnormality and Tau phosphorylation, with 11 FDR-significant proteins; Aβ PET positivity also rose from 10.3% to 89.8% across this boundary. Later differences were more selective. GFAP differed across all four adjacent-stage comparisons, whereas NfL showed its largest difference between Moderate aggregation and Advanced aggregation (Fig. 4b and Supplementary Table S5). Rule-based exemplar profiles illustrated these patterns (Fig. 4c).

**Figure 4.**
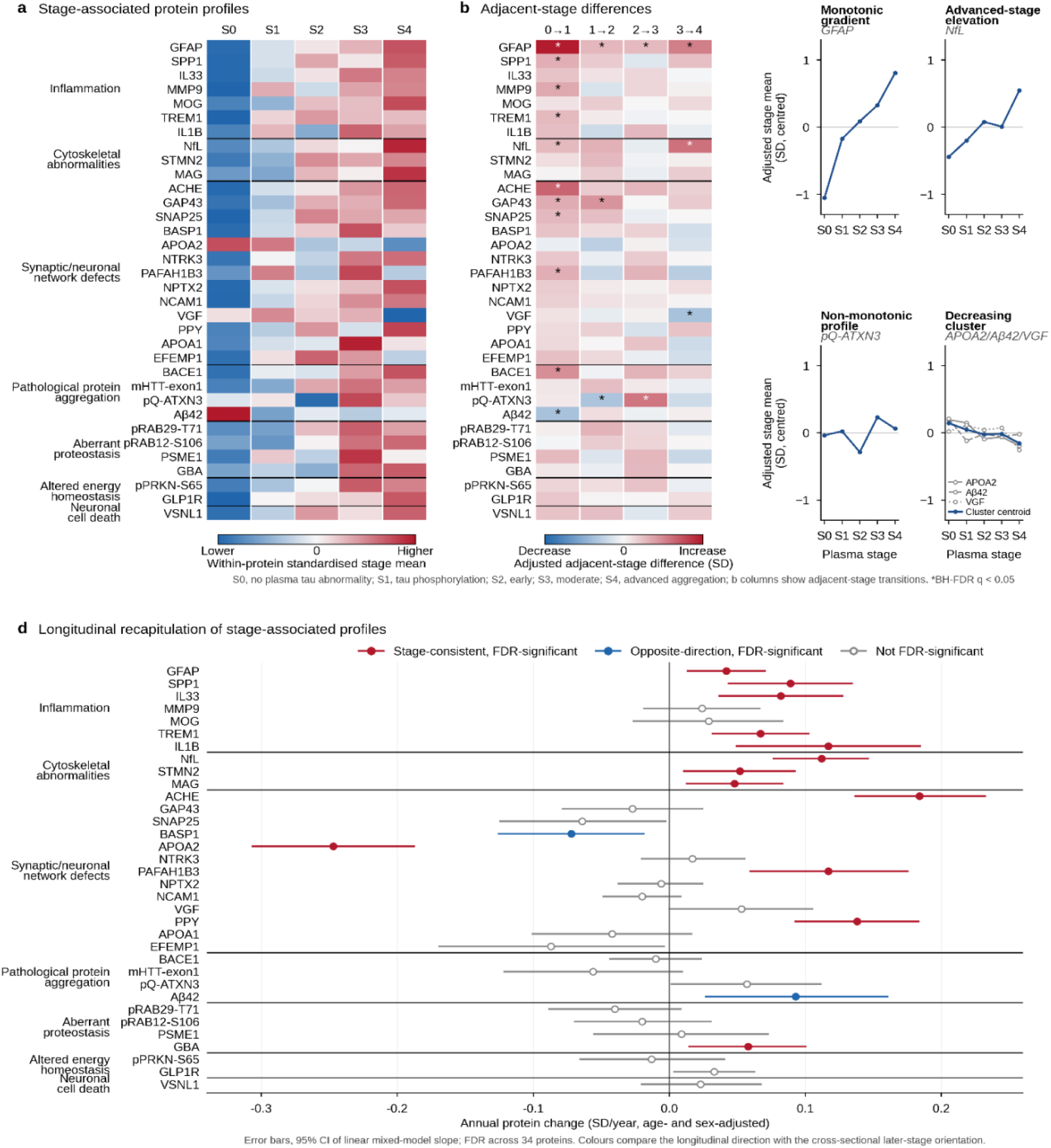
Biological characterisation and longitudinal recapitulation. The same 34 stage-associated non-tau proteins are shown. (a) Age-and sex-adjusted stage profiles. (b) Adjacent-stage differences; asterisks indicate BH-FDR q<0.05 across 136 contrasts. (c) Rule-based exemplars illustrate the GFAP gradient, advanced-stage NfL elevation, non-monotonic pQ-ATXN3 profile and complete decreasing cluster (APOA2, Aβ42 and VGF). (d) Annual protein change with 95% CIs in 306 participants with 640 draws; proteins are ordered as in panels a and b and grouped by the same predefined Neuro220 domains. Red, blue and open symbols denote FDR-significant stage-consistent, FDR-significant opposite-direction and nonsignificant changes, respectively. Fifteen proteins changed, 13 in the stage-consistent direction. Adjacent-stage comparisons are cross-sectional. Aβ, amyloid-β; BH-FDR, Benjamini–Hochberg false discovery rate; CI, confidence interval; GFAP, glial fibrillary acidic protein; NfL, neurofilament light chain.

The same 34 proteins were evaluated in the serial nucleic acid-linked immuno-sandwich assay (NULISA) cohort (306 participants; 640 blood draws). Fifteen changed longitudinally after BH-FDR correction, and 13 of 15 changed in the stage-consistent direction (Fig. 4d, Supplementary Fig. S3 and Supplementary Table S5). Longitudinally changing proteins included GFAP, SPP1, triggering receptor expressed on myeloid cells 1 (TREM1), interleukin-33 (IL33), interleukin-1β (IL1B), NfL, stathmin-2 (STMN2) and myelin-associated glycoprotein (MAG).

## Discussion

In this multicentre observational study, we used a single high-plex immunoassay platform to directly compare prespecified T1/T2 assays and multianalyte models and to build a plasma biological staging framework aligned with the revised Core 1/T1–Core 2/T2 architecture. Serial clinical and proteomic measurements were then used to validate the hierarchy and characterise the broader biology associated with each stage. Our major findings were as follows. First, BD-p-tau217 and eMTBR-tau were selected as the most informative T1 and T2 biomarkers, respectively, whereas multianalyte alternatives did not improve discrimination. Second, the five-stage hierarchy distinguished longitudinal CDR-SB trajectories across all four adjacent-stage contrasts, and forward within-person stage movement was associated with faster clinical worsening than stage stability. Finally, the plasma-defined stages mapped onto broader non-tau proteomic patterns, with transition-specific remodelling and partial longitudinal recapitulation in serial samples. Taken together, our findings suggest that complementary T1 and T2 plasma biomarkers define disease position, whereas high-plex profiling reveals the broader, partly dynamic biological state accompanying that position. This separation could support scalable stage assignment while providing a framework for stage-matched biological phenotyping and for prioritising dynamic pathways in longitudinal monitoring and therapeutic research.

Our first major finding was that BD-p-tau217 and eMTBR-tau emerged as the most informative single-marker representations of the T1 and T2 axes, respectively, forming a parsimonious two-marker backbone. Recent studies established the temporal ordering of plasma biomarkers spanning these axes, the sequential diagnostic use of p-tau217 and eMTBR-tau, and their ability to approximate PET-based biological stages.^7–11^ However, they did not systematically compare the broader candidate landscape of conventional and brain-derived assays and multianalyte representations within a single analytical platform. In our head-to-head analyses, the four-marker BD T1 composite performed slightly worse than BD-p-tau217 alone, while adding BD-p-tau205 provided no gain at the early or moderate tau PET anchors and reduced discrimination at the advanced tau PET anchor. Biologically, BD-p-tau217 functioned as an amyloid-associated, Core 1-aligned T1 signal, whereas eMTBR-tau functioned as a later Core 2-aligned measure of established AD tau proteinopathy. Thus, the value of the high-plex platform in building the framework was not to maximise marker number, but to identify the simplest robust T1/T2 representation while preserving the remaining proteome for independent biological characterisation.

Our second major finding was that the five-stage hierarchy was clinically ordered both between and within individuals: it distinguished longitudinal CDR-SB trajectories across all four adjacent-stage boundaries, and forward stage movement was coupled to faster clinical worsening than stage stability. The between-stage gradient is consistent with previous PET-and plasma-based staging studies linking higher baseline stage to greater clinical severity and faster subsequent progression. The more distinctive contribution was the within-person analysis. By reapplying the prespecified staging algorithm at serial plasma measurements, we showed that forward movement through the hierarchy predominated and was accompanied by faster CDR-SB worsening. These findings extend plasma staging beyond static disease-position assignment and support a scalable framework for monitoring biological progression and its clinical relevance over time.

Our final major finding was that plasma-defined stages mapped onto broader non-tau proteomic patterns, with transition-specific remodelling that was partly recapitulated longitudinally. Previous plasma proteomic studies have identified coordinated pathway changes across the AD continuum, but have generally not localised these across ordered plasma-defined stage boundaries. Screening all 197 eligible non-tau proteins identified 34 stage-associated proteins spanning multiple neurodegenerative pathways. The composition of this signature was biologically coherent with processes implicated in AD,^22–26^ encompassing inflammatory and immune-related proteins such as SPP1, TREM1 and MMP9; synaptic-neuronal proteins including ACHE, GAP43 and SNAP25; amyloid-processing signals including BACE1 and Aβ42; and aggregation-or proteostasis-related proteins such as pQ-ATXN3 and GBA. Adjacent-stage analyses further showed that this remodelling was not uniform across the hierarchy. The transition to an amyloid-associated T1-positive state coincided with the broadest proteomic shift, whereas later T2-positive aggregation stages showed more selective remodelling: astroglial and synaptic changes at entry into Early aggregation; astroglial and aggregation-related changes at entry into Moderate aggregation; and astroglial, neuroaxonal and synaptic changes at entry into Advanced aggregation. This stage localisation was also biologically coherent with the revised criteria: GFAP differed across all four adjacent-stage boundaries, consistent with sustained I-related astrocytic reactivity, whereas NfL showed its largest difference on entry into Advanced aggregation, consistent with greater N-related neuroaxonal injury. Together, this transition-resolved map indicates that plasma-defined disease position organises broader stage-associated biology that cannot be inferred from the defining T1/T2 biomarkers alone and provides a basis for stage-matched biological phenotyping.

More importantly, serial high-plex profiling provided a within-person test of the cross-sectional stage map. Fifteen of the 34 stage-associated proteins changed longitudinally after false-discovery-rate correction, and 13 moved in the direction predicted by their later-stage cross-sectional profiles. This recapitulation was biologically structured rather than driven by a single marker: GFAP was consistent with persistent astrocytic reactivity; SPP1, IL33, TREM1 and IL1B with inflammatory and innate immune activity; and NfL, STMN2 and MAG with axonal, cytoskeletal and myelin-related injury. Stage-consistent changes also extended to proteins assigned to synaptic-neuronal processes (ACHE, APOA2, PAFAH1B3 and PPY) and proteostasis (GBA). Consistent with these protein-level findings, inflammation and cytoskeletal domain scores increased longitudinally. The convergence of cross-sectional stage ordering, within-person protein change and coherent domain-level shifts suggests that part of the stage-associated proteomic architecture reflects active biological remodelling rather than stable between-person differences alone. These longitudinally responsive proteins and pathways provide a biologically prioritised set of candidates for stage-matched monitoring and future mechanistic or therapeutic studies.^27,28^

Strengths include multicentre recruitment, head-to-head evaluation of prespecified T1/T2 assays and multianalyte models within a common high-plex platform, and serial clinical and proteomic measurements. Several limitations should be considered. First, although the regional tau PET anchors were defined in a broader Samsung Medical Center (SMC) dataset, the plasma eMTBR-tau thresholds were derived and evaluated in the same 116-participant plasma–tau PET sample; independent external calibration is therefore required, and the NULISA Protein Quantification (NPQ) cutoffs remain platform-and workflow-specific. Second, the adjacent-stage proteomic contrasts were cross-sectional, follow-up duration and the number of repeated samples varied, and some participants showed reverse or unstageable transitions; longitudinal recapitulation therefore supports, but does not establish, a fixed temporal sequence for every protein or participant. Finally, the high-plex panel was a targeted plasma panel rather than an unbiased representation of brain biology; circulating proteins cannot establish cellular source or causality, multiple exploratory analysis families remained despite FDR control, and the predominantly Korean cohort may limit generalisability across ancestries and clinical settings.^29^ Nevertheless, the convergence of PET-linked marker selection, adjacent-stage clinical gradients, within-person stage movement and serial proteomic recapitulation provides internally consistent support for the ordered framework.

In conclusion, a parsimonious plasma backbone combining a Core 1-aligned T1 biomarker and a Core 2-aligned T2 biomarker defined an ordered disease-position hierarchy supported by longitudinal clinical trajectories and within-person stage movement. High-plex profiling showed that these plasma-defined positions corresponded to broader, partly dynamic biological states, providing a framework for stage-matched phenotyping and future longitudinal biomarker studies.

## Methods

### Study design and participants

The Korea-Registries to Overcome Dementia and Accelerate Dementia Research (K-ROAD) is a prospective multicentre registry with standardised clinical assessment, neuropsychological testing, brain imaging and biobanking.^30^ The parent NULISA Neuro220 Panel dataset comprised 1,394 participants and 2,034 blood draws; 1,035 participants with baseline NULISAseq Neuro 220 measurements and the data required for stage construction or validation formed the baseline staging cohort (Fig. 1). Participants were classified as cognitively unimpaired, mild cognitive impairment or dementia by multidisciplinary assessment. Stage construction used baseline data. Longitudinal clinical and proteomic analyses were restricted to participants untreated with anti-amyloid disease-modifying therapy during the relevant observation period; symptomatic treatments were permitted.

The study was approved by the institutional review boards of participating centres, and all participants or their legally authorised representatives provided written informed consent. Reporting followed the STROBE guideline, with STARD items applied to the diagnostic-accuracy analyses.^31,32^

### Plasma proteomic measurements

EDTA plasma was processed using standardised procedures, stored at −80 °C and analysed with the proximity-based NULISAseq Neuro 220 Panel (Alamar Biosciences, Fremont, CA, USA).^15,17^ Manufacturer-normalised NULISA Protein Quantification (NPQ) values were reported on a log2 scale. Samples were assayed in two phases, with longitudinal samples from the same participant kept together whenever possible. Prespecified sample, internal, inter-plate and negative-control criteria were applied. After exclusion of poorly detected targets and the APOE4 protein analyte, 210 analytes were available for continuous analyses. NPQ values were corrected for assay phase and batch before analysis; these technical factors were not included again as covariates in the statistical models.

### Imaging and clinical assessments

Aβ PET was acquired with [18F]florbetaben or [18F]flutemetamol, co-registered to individual MRI and quantified in Centiloid units using BeauBrain Amylo software.^33–35^ Aβ PET positivity was defined as Centiloid >24.

Tau PET was acquired with [18F]flortaucipir and regional standardised uptake value ratios were calculated using inferior cerebellar grey matter as the reference region without partial-volume correction.^36^ FreeSurfer version 6.0 was used for native-space parcellation.^37^ The Temporal Meta region comprised the entorhinal cortex, amygdala, fusiform gyrus, parahippocampal gyrus, inferior temporal cortex and lingual gyrus; the neocortical composite (NeoC) excluded the precentral, postcentral and paracentral gyri. Regional tau PET measurements were available in 116 participants with matched baseline plasma; when multiple scans were available, the scan closest to blood sampling was used. Three tau PET anchors, fixed from the full SMC tau PET dataset (n=455) before the plasma T2 analyses, were applied: Temporal Meta >1.376, NeoC >1.248 and high NeoC >1.6015. The Temporal Meta and NeoC positivity thresholds were defined as the mean plus 2 SD among Aβ PET-negative cognitively unimpaired participants. The high-NeoC threshold was set at the 66.67th percentile of NeoC SUVR among tau PET-positive participants, corresponding to the upper-tertile boundary. Tau PET positivity for this derivation was defined as positivity in any of the medial temporal lobe, Temporal Meta or neotemporal regions.

Clinical severity was assessed using the Clinical Dementia Rating–Sum of Boxes (CDR-SB), and global cognition was described with the Mini-Mental State Examination (MMSE).^38,39^ For longitudinal CDR-SB analyses, time zero was the baseline blood draw; assessments obtained within 1 year before baseline and all subsequent assessments were retained, with time expressed in years from baseline.

### Building the plasma biological staging framework

T1 was prespecified as a Core 1-aligned biomarker axis of amyloid-associated phosphorylated and secreted AD tau, whereas T2 was prespecified as a Core 2-aligned axis of established AD tau proteinopathy.^1,7–11^ T1 candidates were p-tau181, p-tau212, p-tau217 and p-tau231, together with brain-derived (BD) assays targeting the same phosphosites where available; T2 candidates were p-tau205, BD-p-tau205 and eMTBR-tau243. Assays targeting the same phosphosite were compared head-to-head. Clinical outcomes and the broader non-tau proteome were withheld from marker selection and threshold derivation.

T1 candidates were benchmarked against Aβ PET positivity and continuous Centiloid, and T2 candidates against the three nested tau PET anchors. The best-performing single biomarker for each axis was selected, and thresholds were derived using Youden’s J statistic with bootstrap assessment of stability. Prespecified multianalyte sensitivity models tested whether combining candidates improved performance. The selected biomarkers and thresholds were fixed before clinical and longitudinal validation and broader biological characterisation. Stage assignment used the selected T1 threshold and ordered T2 thresholds; T1-negative/T2-positive profiles were classified as unstageable rather than forced into the hierarchy.

### Statistical analysis

Continuous variables are summarised as mean (SD) or median (IQR), as appropriate, and categorical variables as n (%). Candidate-biomarker performance was evaluated using area under the receiver-operating-characteristic curve (AUC), Spearman correlation with continuous PET burden, paired bootstrap comparisons between assay formats^40^ and multianalyte sensitivity models. Threshold stability was assessed by bootstrap resampling. Baseline ordinal stage and CDR-SB were related using Spearman correlation.

For longitudinal clinical validation, linear mixed-effects models included time, baseline stage and stage-by-time interaction, with participant-specific random intercepts and slopes. The global interaction was tested by likelihood-ratio comparison, and four prespecified adjacent-stage slope contrasts were controlled using the Benjamini–Hochberg false-discovery-rate (BH-FDR) procedure.^41^ In the serial cohort, frozen thresholds were reapplied at each blood draw and first-to-last transitions were classified as progression, stability, regression or unstageable at follow-up. Directional asymmetry was tested with an exact binomial test. Clinical coupling was evaluated among participants stageable at both draws with baseline Stage 0–3 and eligible longitudinal CDR-SB assessments using mixed-effects models containing time-by-transition-group and time-by-baseline-stage interactions. Progression-versus-stable and regression-versus-stable contrasts were BH-FDR controlled. Because blood-transition and CDR-SB observation windows overlapped, this analysis was interpreted as longitudinal coupling rather than landmark prediction.

For broader proteomic characterisation, the complete tau/T1/T2/MAPT family was further excluded from the 210 continuous analytes, leaving 197 non-tau proteins. Each standardised protein was modelled as a function of categorical stage with adjustment for age and sex. Four-degree-of-freedom omnibus tests were BH-FDR controlled across 197 proteins; partial R² quantified incremental variance explained by stage. Secondary ordinal-stage models used heteroscedasticity-robust standard errors. For FDR-positive proteins, four age-and sex-adjusted adjacent-stage contrasts were estimated with robust 95% CIs and BH-FDR correction across the complete contrast family. Manufacturer-defined Neuro220 domains were used only after protein selection for biological annotation.

To test longitudinal recapitulation of the cross-sectional stage pattern, annual change in the same FDR-positive proteins was estimated using separate linear mixed-effects models with participant-specific random intercepts and slopes. Cross-sectional direction was fixed before longitudinal testing, and BH-FDR correction was applied across proteins. Equal-weight stage-oriented domain scores were analysed for predefined domains containing at least three selected proteins, and directional concordance was assessed among proteins changing longitudinally after FDR correction.

All tests were two-sided, 95% CIs were used, and analyses were performed in Python 3.10 with statsmodels and SciPy.^42,43^

## Data availability

Deidentified individual participant data are not publicly available because of participant privacy and institutional data-governance restrictions. Data may be made available to qualified researchers upon reasonable request to the corresponding authors, subject to approval by the relevant institutional committees and execution of applicable data-use agreements.

## Code availability

Analysis code supporting the findings of this study is available from the corresponding authors upon reasonable request, subject to institutional and data-governance requirements.

## Acknowledgements

We thank the participants and their families and acknowledge the investigators and staff of the K-ROAD study group. We also acknowledge the Alamar Biosciences Technology Access Program for support of NULISAseq Neuro 220 profiling.

## Author contributions

Yuna Gu contributed to conceptualization, methodology, formal analysis, software, visualization, data curation, writing of the original draft, and review and editing of the manuscript. Jun Pyo Kim contributed to conceptualization, methodology, formal analysis, data curation, project administration, writing of the original draft, and review and editing of the manuscript. Bo Hyun Kim, Hyunwoo Lee, Jungah Lee, Heekyoung Kang, Sohyun Yim, Daeun Shin, Hee Kyung Park, and Seonghyeon Kim contributed to investigation, data curation, and review and editing. Byung Hwa Lee, Jeongha Lee, and Hyeongjin Kim contributed to investigation and resources. Henrik Zetterberg contributed to review and editing. Min Young Chun, Duk L. Na, Jae-Hong Lee, Soo Hyun Cho, Byeong C. Kim, Si Eun Kim, Gyeongmo Sohn, Jihwan Yun, Jae-Sung Lim, Juyoung Kim, Kyungbok Lee, and Jaeho Kim contributed to investigation, resources, data curation, and review and editing. Hee Jin Kim and Eun-Joo Kim contributed to conceptualization, and review and editing. Hyemin Jang and Sang Won Seo contributed to conceptualization, supervision, funding acquisition, project administration, and review and editing.

## References

1. Jack Jr, C.R., et al. Revised criteria for diagnosis and staging of Alzheimer’s disease: Alzheimer’s Association Workgroup. Alzheimer’s & Dementia 20, 5143–5169 (2024).

2. Pichet Binette, A., et al. Evaluation of the revised criteria for biological and clinical staging of Alzheimer disease. JAMA neurology 82, 666–675 (2025).

3. Therriault, J., et al. Biomarker modeling of Alzheimer’s disease using PET-based Braak staging. Nature aging 2, 526–535 (2022).

4. Therriault, J., et al. Biomarker-based staging of Alzheimer disease: rationale and clinical applications. Nature Reviews Neurology 20, 232–244 (2024).

5. Palmqvist, S., et al. Blood biomarkers to detect Alzheimer disease in primary care and secondary care. Jama 332, 1245–1257 (2024).

6. Palmqvist, S., et al. Alzheimer’s Association Clinical Practice Guideline on the use of blood-based biomarkers in the diagnostic workup of suspected Alzheimer’s disease within specialized care settings. Alzheimer’s & Dementia 21, e70535 (2025).

7. Collij, L.E., et al. Trajectories of plasma and CSF MTBR-tau243 and phosphorylated-tau species across the Alzheimer’s disease continuum. Nature Communications 17, 3400 (2026).

8. Horie, K., et al. Plasma MTBR-tau243 biomarker identifies tau tangle pathology in Alzheimer’s disease. Nature medicine 31, 2044–2053 (2025).

9. Mattsson-Carlgren, N., et al. Integration of plasma eMTBR-tau243 and p-tau217 in the diagnosis and stratification of Alzheimer’s disease: a prospective cohort study. The Lancet Neurology 25, 357–367 (2026).

10. Montoliu-Gaya, L., et al. Plasma tau biomarkers for biological staging of Alzheimer’s disease. Nature Aging 5, 2297–2308 (2025).

11. Salvadó, G., et al. Plasma eMTBR-tau243 and %p-tau217 for biological staging of Alzheimer disease. JAMA neurology (2026).

12. Barthélemy, N.R., et al. Highly accurate blood test for Alzheimer’s disease is similar or superior to clinical cerebrospinal fluid tests. Nature medicine 30, 1085–1095 (2024).

13. Gonzalez-Ortiz, F., et al. Brain-derived tau: a novel blood-based biomarker for Alzheimer’s disease-type neurodegeneration. Brain 146, 1152–1165 (2023).

14. Warmenhoven, N., et al. A comprehensive head-to-head comparison of key plasma phosphorylated tau 217 biomarker tests. Brain 148, 416–431 (2025).

15. Ashton, N.J., et al. Biomarker discovery in Alzheimer’s and neurodegenerative diseases using Nucleic Acid Linked Immuno-Sandwich Assay. Alzheimer’s & Dementia 21, e14621 (2025).

16. Bellomo, G., et al. Plasma proteome profiling identified biomarkers for the differential diagnosis and molecular staging of neurodegenerative dementias. Nature Aging, 1–16 (2026).

17. Feng, W., et al. NULISA: a proteomic liquid biopsy platform with attomolar sensitivity and high multiplexing. Nature communications 14, 7238 (2023).

18. Guo, Y., et al. Plasma proteomic profiles predict future dementia in healthy adults. Nature aging 4, 247–260 (2024).

19. Johnson, E.C., et al. Large-scale proteomic analysis of Alzheimer’s disease brain and cerebrospinal fluid reveals early changes in energy metabolism associated with microglia and astrocyte activation. Nature medicine 26, 769–780 (2020).

20. Johnson, E.C.B., et al. Cerebrospinal fluid proteomics define the natural history of autosomal dominant Alzheimer’s disease. Nature Medicine 29, 1979–1988 (2023).

21. Vogel, J.W., et al. Four distinct trajectories of tau deposition identified in Alzheimer’s disease. Nature medicine 27, 871–881 (2021).

22. De Schepper, S., et al. Perivascular cells induce microglial phagocytic states and synaptic engulfment via SPP1 in mouse models of Alzheimer’s disease. Nat. Neurosci. 26, 406–415 (2023).

23. Khalil, M., et al. Neurofilaments as biomarkers in neurological disorders. Nature Reviews Neurology 14, 577–589 (2018).

24. Pereira, J.B., et al. Plasma GFAP is an early marker of amyloid-β but not tau pathology in Alzheimer’s disease. Brain 144, 3505–3516 (2021).

25. Schindler, S.E., et al. High-precision plasma β-amyloid 42/40 predicts current and future brain amyloidosis. Neurology 93, e1647–e1659 (2019).

26. Wilson, E.N., et al. TREM1 disrupts myeloid bioenergetics and cognitive function in aging and Alzheimer disease mouse models. Nat. Neurosci. 27, 873–885 (2024).

27. Sims, J.R., et al. Donanemab in early symptomatic Alzheimer disease: the TRAILBLAZER-ALZ 2 randomized clinical trial. JAMA 330, 512–527 (2023).

28. van Dyck, C.H., et al. Lecanemab in early Alzheimer’s disease. N. Engl. J. Med. 388, 9–21 (2023).

29. Lin, Y.-S., et al. Cross-cultural validation of plasma p-tau217 and p-tau181 as precision biomarkers for amyloid PET positivity: an East Asian study in Taiwan and Korea. Alzheimer’s & Dementia 21, e14565 (2025).

30. Jang, H., et al. Korea-Registries to Overcome Dementia and Accelerate Dementia Research (K-ROAD): a cohort for dementia research and ethnic-specific insights. Dementia and Neurocognitive Disorders 23, 212 (2024).

31. Bossuyt, P.M., et al. STARD 2015: an updated list of essential items for reporting diagnostic accuracy studies. Radiology 277, 826–832 (2015).

32. Von Elm, E., et al. The Strengthening the Reporting of Observational Studies in Epidemiology (STROBE) statement: guidelines for reporting observational studies. The lancet 370, 1453–1457 (2007).

33. Cho, S.H., et al. A new Centiloid method for 18F-florbetaben and 18F-flutemetamol PET without conversion to PiB. European Journal of Nuclear Medicine and Molecular Imaging 47, 1938–1948 (2020).

34. Kim, S.-J., et al. Development and clinical validation of CT-based regional modified Centiloid method for amyloid PET. Alzheimer’s Research & Therapy 14, 157 (2022).

35. Klunk, W.E., et al. The Centiloid Project: standardizing quantitative amyloid plaque estimation by PET. Alzheimer’s & dementia 11, 1–15. e14 (2015).

36. Maass, A., et al. Comparison of multiple tau-PET measures as biomarkers in aging and Alzheimer’s disease. Neuroimage 157, 448–463 (2017).

37. Fischl, B. FreeSurfer. Neuroimage 62, 774–781 (2012).

38. Folstein, M.F., Folstein, S.E. & McHugh, P.R. “Mini-mental state”: a practical method for grading the cognitive state of patients for the clinician. Journal of psychiatric research 12, 189–198 (1975).

39. Morris, J.C. The Clinical Dementia Rating (CDR) current version and scoring rules. Neurology 43, 2412–2412-a (1993).

40. DeLong, E.R., DeLong, D.M. & Clarke-Pearson, D.L. Comparing the areas under two or more correlated receiver operating characteristic curves: a nonparametric approach. Biometrics 44, 837–845 (1988).

41. Benjamini, Y. & Hochberg, Y. Controlling the false discovery rate: a practical and powerful approach to multiple testing. J. R. Stat. Soc. B 57, 289–300 (1995).

42. Seabold, S. & Perktold, J. Statsmodels: econometric and statistical modeling with python. scipy 7, 92–96 (2010).

43. Virtanen, P., et al. SciPy 1.0: fundamental algorithms for scientific computing in Python. Nature methods 17, 261–272 (2020).

