## Supplementary Information for "Plasma proteomics reveals stage-dependent biological remodelling in Alzheimer’s disease"

**Supplementary Appendix**

Supplementary Figures S1–S3 and Supplementary Tables S1–S5

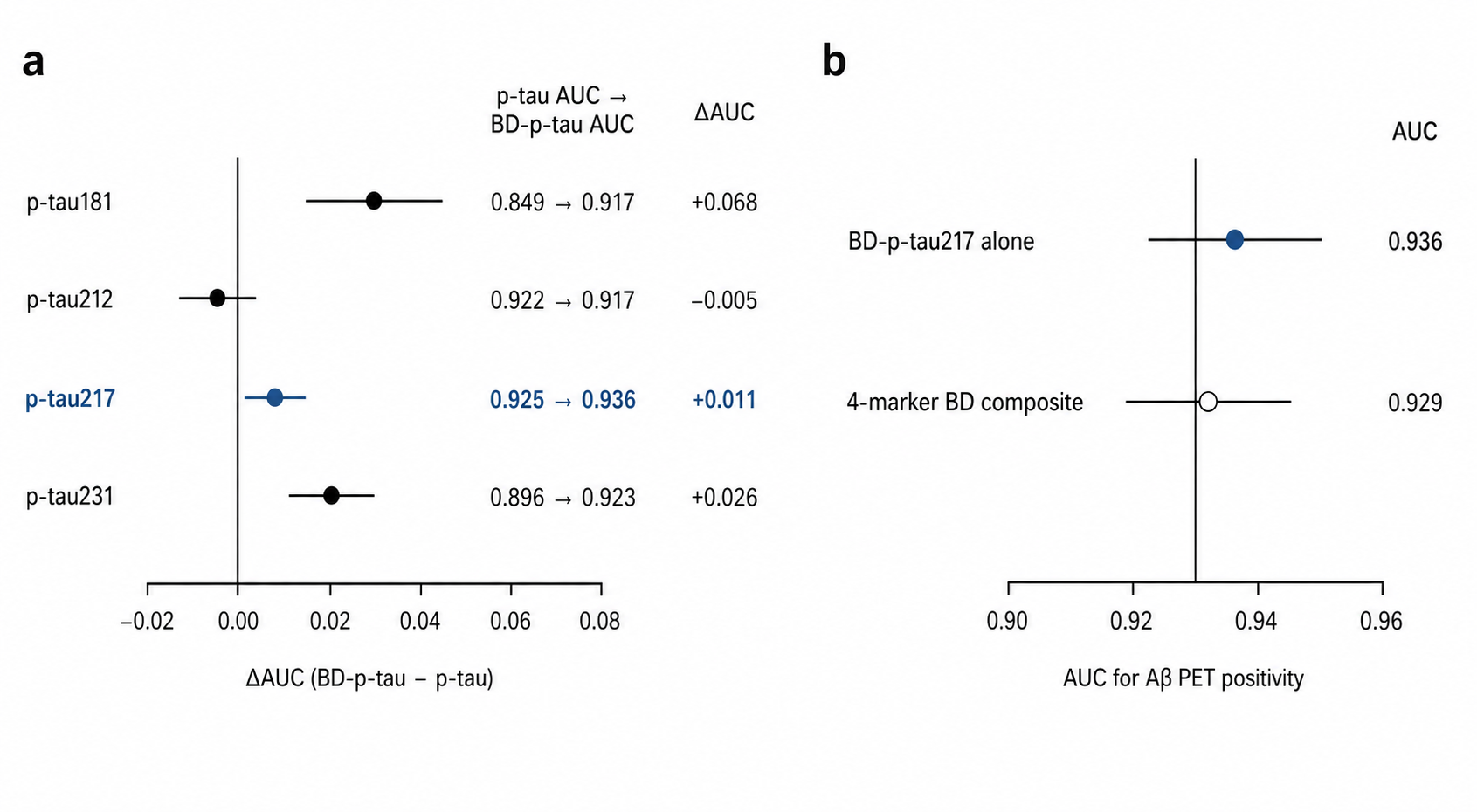

Supplementary Figure S1. Selection of BD-p-tau217 as the T1 plasma biomarker. (A) Head-to-head comparison of p-tau and brain-derived p-tau (BD-p-tau) assays for p-tau181, p-tau212, p-tau217, and p-tau231. Points show paired differences in AUC (BD-p-tau minus p-tau) with bootstrap 95% CIs; adjacent text shows p-tau AUC to BD-p-tau AUC. (B) Comparison of BD-p-tau217 alone with an equal-weight four-marker BD composite comprising BD-p-tau181, BD-p-tau212, BD-p-tau217, and BD-p-tau231. The composite did not improve discrimination of Aβ PET positivity beyond BD-p-tau217 alone. Aβ PET positivity was defined as Centiloid >24. AUC, area under the receiver-operating-characteristic curve; BD, brain-derived; CI, confidence interval; PET, positron emission tomography.

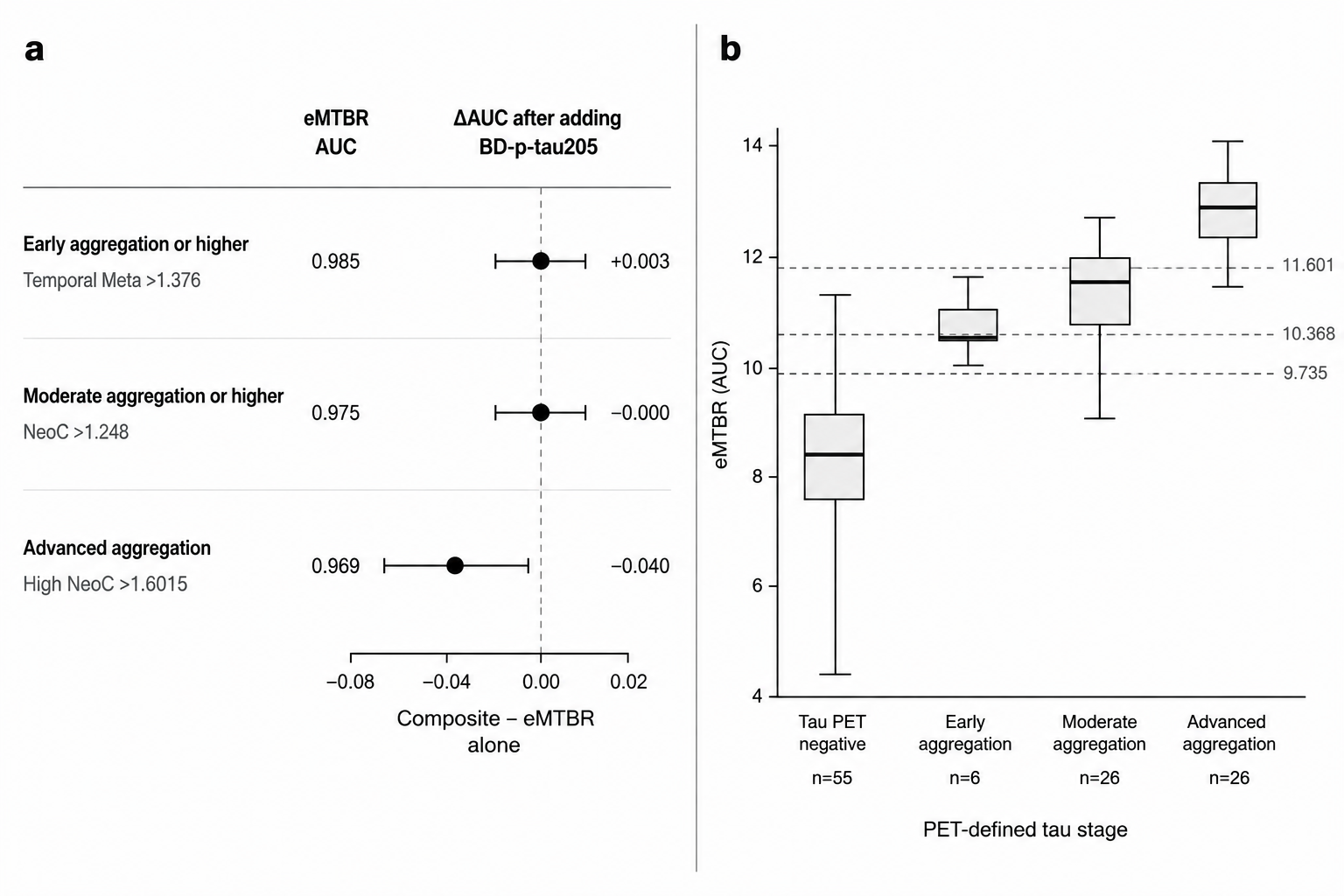

**Supplementary Figure S2.** T2 marker-selection robustness. (A) Comparison of eMTBR-tau243 alone with a composite incorporating BD-p-tau205 across the prespecified tau PET anchors. Points show the change in AUC after adding BD-p-tau205, with bootstrap 95% CIs; the composite did not provide consistent incremental discrimination beyond eMTBR-tau243 alone. (B) Distribution of eMTBR-tau243 across the final PET-defined tau hierarchy: tau PET negative (n=55), early aggregation (n=9), moderate aggregation (n=26), and advanced aggregation (n=26). Dashed lines indicate the frozen eMTBR-tau243 thresholds of 9.735, 10.358, and 11.608 log2 NPQ. BD, brain-derived; CI, confidence interval; eMTBR-tau243, endogenously cleaved microtubule-binding-region tau243; NPQ, NULISA Protein Quantification; PET, positron emission tomography.

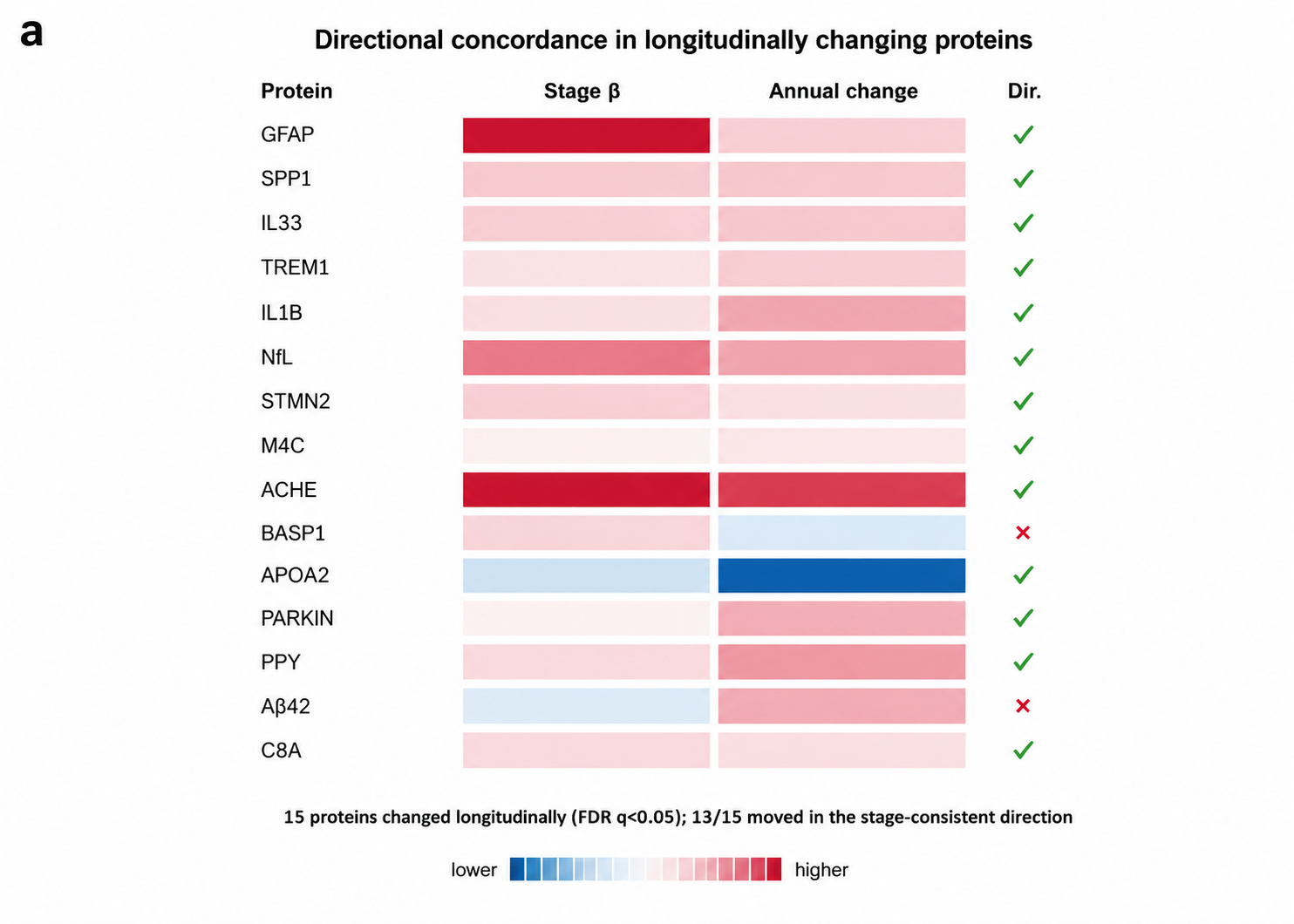

Supplementary Figure S3. Longitudinal proteomic recapitulation. Directional concordance between the cross-sectional ordinal stage effect and longitudinal change among proteins that changed after BH-FDR correction. Fifteen proteins changed longitudinally, and 13 of 15 moved in the stage-consistent direction. BH-FDR, Benjamini-Hochberg false discovery rate.

**Supplementary Table S1. T1 candidate assay performance and marker-selection robustness**

**A. Candidate T1 assays benchmarked against Aβ PET**

| **Assay** | **N** | **Missing** | **AUC for Aβ PET positivity** | **95% CI** | **Spearman ρ vs Centiloid** | **95% CI** |
| --- | --- | --- | --- | --- | --- | --- |
| BD-p-tau181 | 1,035 | 0 | 0.917 | 0.895–0.939 | 0.676 | 0.642–0.711 |
| p-tau181 | 1,035 | 0 | 0.849 | 0.821–0.877 | 0.572 | 0.525–0.614 |
| BD-p-tau212 | 1,035 | 0 | 0.917 | 0.894–0.939 | 0.681 | 0.645–0.717 |
| p-tau212 | 1,035 | 0 | 0.922 | 0.899–0.942 | 0.688 | 0.649–0.722 |
| BD-p-tau217 | 1,035 | 0 | 0.936 | 0.916–0.956 | 0.706 | 0.671–0.739 |
| p-tau217 | 1,035 | 0 | 0.925 | 0.903–0.945 | 0.692 | 0.656–0.725 |
| BD-p-tau231 | 1,035 | 0 | 0.923 | 0.901–0.942 | 0.687 | 0.652–0.723 |
| p-tau231 | 1,035 | 0 | 0.896 | 0.872–0.920 | 0.645 | 0.602–0.683 |

B. Paired p-tau-versus-BD-p-tau assay comparison

| **Phosphosite** | Assay-pair ρ | BD-p-tau AUC | p-tau AUC | ΔAUC (BD-p-tau−p-tau) | **95% CI** | **Bootstrap P** |
| --- | --- | --- | --- | --- | --- | --- |
| p-tau181 | 0.876 | 0.917 | 0.849 | 0.068 | 0.051–0.086 | <0.001 |
| p-tau212 | 0.946 | 0.917 | 0.922 | −0.005 | −0.017–0.006 | 0.445 |
| p-tau217 | 0.951 | 0.936 | 0.925 | 0.011 | 0.004–0.018 | 0.003 |
| p-tau231 | 0.944 | 0.923 | 0.896 | 0.026 | 0.015–0.038 | <0.001 |

**C. Single-marker versus multianalyte T1 representations**

| **T1 representation** | **AUC** | **95% CI** | **Spearman ρ** | **ΔAUC: BD-p-tau217 − model** | **95% CI** | **Bootstrap P** |
| --- | --- | --- | --- | --- | --- | --- |
| BD-p-tau217 alone | 0.936 | 0.917–0.955 | 0.706 | 0.000 | 0.000–0.000 | 1.000 |
| BD canonical (181+217+231) | 0.930 | 0.908–0.950 | 0.697 | 0.006 | 0.002–0.011 | <0.001 |
| BD expanded (+212) | 0.929 | 0.908–0.949 | 0.698 | 0.007 | 0.003–0.012 | <0.001 |
| p-tau canonical (181+217+231) | 0.902 | 0.878–0.924 | 0.654 | 0.035 | 0.022–0.048 | <0.001 |
| p-tau expanded (+212) | 0.909 | 0.886–0.931 | 0.668 | 0.027 | 0.018–0.038 | <0.001 |
| Site-consensus canonical | 0.921 | 0.898–0.941 | 0.685 | 0.016 | 0.009–0.023 | <0.001 |
| Site-consensus expanded (+212) | 0.923 | 0.901–0.944 | 0.690 | 0.013 | 0.007–0.020 | <0.001 |
| p-tau217 site-consensus | 0.934 | 0.912–0.953 | 0.705 | 0.003 | −2.93×10⁻⁴–0.006 | 0.073 |

**D. Frozen T1 threshold**

| **Population** | **N** | **Aβ+ N** | **BD-p-tau217 cutoff** | **Bootstrap 95% CI** | **Sensitivity** | **Specificity** | **AUC** |
| --- | --- | --- | --- | --- | --- | --- | --- |
| Overall | 1,035 | 692 | 9.563 | 9.346–9.948 | 0.941 | 0.869 | 0.936 |

Aβ PET positivity was defined as Centiloid >24. All T1 candidates were prespecified by biological assignment. The frozen T1 marker was BD-p-tau217; multianalyte representations did not materially improve discrimination beyond the single marker.

**Supplementary Table S2. T2 candidate performance and tau PET anchoring**

**A. Candidate T2 markers against final tau PET anchors (matched n=116)**

| **Tau PET anchor** | **Positive N** | **Marker** | **AUC** | **Youden cutoff** | **Sensitivity** | **Specificity** | **Spearman ρ vs ROI** |
| --- | --- | --- | --- | --- | --- | --- | --- |
| Temporal Meta >1.376 | 61 | p-tau205 | 0.864 | 12.050 | 0.885 | 0.745 | 0.618 |
| Temporal Meta >1.376 | 61 | BD-p-tau205 | 0.943 | 12.660 | 0.902 | 0.836 | 0.791 |
| Temporal Meta >1.376 | 61 | eMTBR-tau243 | 0.986 | 9.735 | 0.984 | 0.945 | 0.870 |
| Neocortical composite >1.248 | 52 | p-tau205 | 0.852 | 12.154 | 0.846 | 0.734 | 0.553 |
| Neocortical composite >1.248 | 52 | BD-p-tau205 | 0.931 | 12.727 | 0.904 | 0.812 | 0.750 |
| Neocortical composite >1.248 | 52 | eMTBR-tau243 | 0.975 | 10.358 | 0.923 | 0.906 | 0.833 |
| High neocortical composite >1.6015 | 26 | p-tau205 | 0.746 | 12.171 | 0.846 | 0.600 | 0.553 |
| High neocortical composite >1.6015 | 26 | BD-p-tau205 | 0.884 | 13.025 | 0.769 | 0.833 | 0.750 |
| High neocortical composite >1.6015 | 26 | eMTBR-tau243 | 0.969 | 11.608 | 0.962 | 0.867 | 0.833 |

**B. Final hierarchical tau PET stages and adjacent-stage discrimination**

| **PET stage** | **Definition** | **N** | p-tau205 adjacent AUC | **BD-p-tau205 adjacent AUC** | **eMTBR-tau243 adjacent AUC** |
| --- | --- | --- | --- | --- | --- |
| Tau PET negative | Temporal Meta ≤1.376 | 55 | — | — | — |
| Early aggregation | Temporal Meta >1.376 and NeoC ≤1.248 | 9 | 0.804 | 0.889 | 0.970 |
| Moderate aggregation | NeoC >1.248 and ≤1.6015 | 26 | 0.735 | 0.709 | 0.778 |
| Advanced aggregation | NeoC >1.6015 | 26 | 0.487 | 0.683 | 0.893 |

Adjacent-stage AUCs compare each row with the immediately preceding PET stage; the tau-PET-negative row therefore has no adjacent AUC. The final T2 marker was eMTBR-tau243. The PET hierarchy was fully nested using Temporal Meta >1.376, neocortical composite >1.248, and high neocortical composite >1.6015. Analyses used full-precision plasma cutoffs of 9.562758948, 9.735217224, 10.35845619 and 11.60847036; manuscript and figure displays show values rounded to three decimals.

Supplementary Table S3. Longitudinal CDR-SB validation according to baseline plasma stage

A. Stage-specific annual CDR-SB slopes in the primary longitudinal clinical-validation cohort

| **Baseline plasma stage** | **N** | **CDR-SB assessments** | **Annual slope, points/year** | **95% CI** |
| --- | --- | --- | --- | --- |
| Stage 0 (No plasma tau abnormality) | 196 | 548 | 0.191 | 0.069–0.313 |
| Stage 1 (Tau phosphorylation) | 128 | 433 | 0.382 | 0.239–0.526 |
| Stage 2 (Early aggregation) | 57 | 188 | 0.674 | 0.464–0.884 |
| Stage 3 (Moderate aggregation) | 101 | 334 | 1.177 | 1.011–1.344 |
| Stage 4 (Advanced aggregation) | 108 | 342 | 1.943 | 1.771–2.114 |

Global stage×time likelihood-ratio χ²(4)=271.386; P=1.60×10⁻⁵⁷. Primary clinical-validation cohort: 590 participants / 1,845 assessments.

**B. Prespecified adjacent-stage slope contrasts**

| **Adjacent-stage contrast** | **Difference in annual slope** | **Nominal P** | **BH-FDR q** |
| --- | --- | --- | --- |
| No plasma tau abnormality → Tau phosphorylation | +0.191 | 0.0466 | 0.0466 |
| Tau phosphorylation → Early aggregation | +0.292 | 0.0247 | 0.0330 |
| Early aggregation → Moderate aggregation | +0.503 | 2.37×10⁻⁴ | 4.73×10⁻⁴ |
| Moderate aggregation → Advanced aggregation | +0.765 | 2.41×10⁻¹⁰ | 9.63×10⁻¹⁰ |

Supplementary Table S4. Proteome-wide cross-sectional association with plasma biological stage

**A. Stage-associated proteins and predefined biological-domain annotation**

| **Protein** | **Predefined Neuro220 domain** | **Partial R²** | **Omnibus q** | **Ordinal β/stage, SD** | **Trend q** |
| --- | --- | --- | --- | --- | --- |
| GFAP | Inflammation | 0.5014 | 3.75e-147 | 0.441 | 5.28e-192 |
| SPP1 | Inflammation | 0.0496 | 7.60e-09 | 0.134 | 1.09e-09 |
| IL33 | Inflammation | 0.0305 | 5.42e-05 | 0.108 | 3.84e-06 |
| MMP9 | Inflammation | 0.0221 | 0.002 | 0.079 | 0.001 |
| MOG | Inflammation | 0.0205 | 0.004 | 0.091 | 2.36e-04 |
| TREM1 | Inflammation | 0.0181 | 0.009 | 0.064 | 0.007 |
| IL1B | Inflammation | 0.0154 | 0.025 | 0.063 | 0.015 |
| NfL | Cytoskeletal abnormalities | 0.1496 | 3.30e-32 | 0.221 | 1.54e-33 |
| STMN2 | Cytoskeletal abnormalities | 0.0257 | 4.69e-04 | 0.097 | 3.61e-05 |
| MAG | Cytoskeletal abnormalities | 0.0137 | 0.048 | 0.071 | 0.006 |
| ACHE | Synaptic and neuronal network defects | 0.2198 | 1.38e-50 | 0.297 | 6.25e-57 |
| GAP43 | Synaptic and neuronal network defects | 0.1230 | 9.94e-26 | 0.224 | 2.74e-26 |
| SNAP25 | Synaptic and neuronal network defects | 0.0371 | 2.90e-06 | 0.113 | 2.39e-06 |
| BASP1 | Synaptic and neuronal network defects | 0.0287 | 1.19e-04 | 0.096 | 1.06e-04 |
| APOA2 | Synaptic and neuronal network defects | 0.0245 | 7.62e-04 | -0.099 | 1.56e-05 |
| NTRK3 | Synaptic and neuronal network defects | 0.0195 | 0.006 | 0.085 | 6.22e-04 |
| PAFAH1B3 | Synaptic and neuronal network defects | 0.0193 | 0.006 | 0.035 | 0.207 |
| NPTX2 | Synaptic and neuronal network defects | 0.0180 | 0.009 | 0.081 | 6.59e-04 |
| NCAM1 | Synaptic and neuronal network defects | 0.0177 | 0.010 | 0.084 | 0.001 |
| VGF | Synaptic and neuronal network defects | 0.0168 | 0.014 | -0.051 | 0.058 |
| PPY | Synaptic and neuronal network defects | 0.0167 | 0.014 | 0.073 | 0.001 |
| APOA1 | Synaptic and neuronal network defects | 0.0143 | 0.038 | 0.062 | 0.025 |
| EFEMP1 | Synaptic and neuronal network defects | 0.0136 | 0.048 | 0.044 | 0.152 |
| BACE1 | Pathological protein aggregation | 0.0976 | 9.68e-20 | 0.195 | 2.86e-20 |
| mHTT-exon1 | Pathological protein aggregation | 0.0228 | 0.002 | 0.092 | 1.18e-04 |
| pQ-ATXN3 | Pathological protein aggregation | 0.0191 | 0.006 | 0.039 | 0.167 |
| Aβ42 | Pathological protein aggregation | 0.0175 | 0.011 | -0.055 | 0.051 |
| pRAB29-T71 | Aberrant proteostasis | 0.0335 | 1.54e-05 | 0.109 | 1.31e-06 |
| pRAB12-S106 | Aberrant proteostasis | 0.0198 | 0.005 | 0.088 | 2.12e-04 |
| PSME1 | Aberrant proteostasis | 0.0195 | 0.006 | 0.063 | 0.011 |
| GBA | Aberrant proteostasis | 0.0153 | 0.026 | 0.076 | 0.002 |
| pPRKN-S65 | Altered energy homeostasis | 0.0330 | 1.81e-05 | 0.114 | 4.23e-07 |
| GLP1R | Altered energy homeostasis | 0.0152 | 0.026 | 0.073 | 0.002 |
| VSNL1 | Neuronal cell death | 0.0405 | 6.39e-07 | 0.124 | 9.31e-08 |

All 197 QC-passed non-tau continuous analytes were tested in the stageable baseline cohort (n=1,007). APOE4 protein analyte and the full tau/T1/T2/MAPT family were excluded before testing. NPQ values were phase/batch-adjusted before analysis. Panel A lists the 34 proteins passing the categorical-stage BH-FDR threshold together with manufacturer-defined Neuro220 domain annotations used only after protein selection. Panel B provides the complete 197-protein results.

**B. Full proteome-wide results for all 197 non-tau proteins**

| **Rank** | **Protein** | **N** | **Partial R²** | **Stage F** | **Omnibus P** | **Omnibus q** | **Ordinal β/stage, SD** | **SE** | **Trend P** | **Trend q** | **Spearman ρ** |
| --- | --- | --- | --- | --- | --- | --- | --- | --- | --- | --- | --- |
| 1 | GFAP | 1007 | 0.5014 | 251.40 | 1.90e-149 | 3.75e-147 | 0.441 | 0.015 | 2.68e-194 | 5.28e-192 | 0.698 |
| 2 | ACHE | 1007 | 0.2198 | 70.43 | 1.40e-52 | 1.38e-50 | 0.297 | 0.018 | 6.35e-59 | 6.25e-57 | 0.425 |
| 3 | NfL | 1007 | 0.1496 | 43.97 | 5.02e-34 | 3.30e-32 | 0.221 | 0.018 | 2.35e-35 | 1.54e-33 | 0.312 |
| 4 | GAP43 | 1007 | 0.1230 | 35.05 | 2.02e-27 | 9.94e-26 | 0.224 | 0.020 | 5.56e-28 | 2.74e-26 | 0.357 |
| 5 | BACE1 | 1007 | 0.0976 | 27.05 | 2.46e-21 | 9.68e-20 | 0.195 | 0.020 | 7.26e-22 | 2.86e-20 | 0.324 |
| 6 | SPP1 | 1007 | 0.0496 | 13.05 | 2.32e-10 | 7.60e-09 | 0.134 | 0.020 | 3.31e-11 | 1.09e-09 | 0.197 |
| 7 | VSNL1 | 1007 | 0.0405 | 10.54 | 2.27e-08 | 6.39e-07 | 0.124 | 0.021 | 3.31e-09 | 9.31e-08 | 0.257 |
| 8 | SNAP25 | 1007 | 0.0371 | 9.65 | 1.18e-07 | 2.90e-06 | 0.113 | 0.021 | 1.22e-07 | 2.39e-06 | 0.207 |
| 9 | pRAB29-T71 | 1007 | 0.0335 | 8.67 | 7.03e-07 | 1.54e-05 | 0.109 | 0.020 | 5.98e-08 | 1.31e-06 | 0.143 |
| 10 | pPRKN-S65 | 1007 | 0.0330 | 8.52 | 9.18e-07 | 1.81e-05 | 0.114 | 0.020 | 1.72e-08 | 4.23e-07 | 0.149 |
| 11 | IL33 | 1007 | 0.0305 | 7.87 | 3.02e-06 | 5.42e-05 | 0.108 | 0.021 | 2.15e-07 | 3.84e-06 | 0.139 |
| 12 | BASP1 | 1007 | 0.0287 | 7.39 | 7.24e-06 | 1.19e-04 | 0.096 | 0.022 | 7.51e-06 | 1.06e-04 | 0.134 |
| 13 | STMN2 | 1007 | 0.0257 | 6.59 | 3.09e-05 | 4.69e-04 | 0.097 | 0.020 | 2.38e-06 | 3.61e-05 | 0.093 |
| 14 | APOA2 | 1007 | 0.0245 | 6.28 | 5.42e-05 | 7.62e-04 | -0.099 | 0.020 | 9.53e-07 | 1.56e-05 | -0.141 |
| 15 | mHTT-exon1 | 1007 | 0.0228 | 5.85 | 1.19e-04 | 0.002 | 0.092 | 0.021 | 8.98e-06 | 1.18e-04 | 0.128 |
| 16 | MMP9 | 1007 | 0.0221 | 5.64 | 1.72e-04 | 0.002 | 0.079 | 0.021 | 1.26e-04 | 0.001 | 0.087 |
| 17 | MOG | 1007 | 0.0205 | 5.22 | 3.63e-04 | 0.004 | 0.091 | 0.021 | 2.04e-05 | 2.36e-04 | 0.095 |
| 18 | pRAB12-S106 | 1007 | 0.0198 | 5.06 | 4.84e-04 | 0.005 | 0.088 | 0.020 | 1.72e-05 | 2.12e-04 | 0.126 |
| 19 | PSME1 | 1007 | 0.0195 | 4.98 | 5.61e-04 | 0.006 | 0.063 | 0.020 | 0.002 | 0.011 | 0.098 |
| 20 | NTRK3 | 1007 | 0.0195 | 4.97 | 5.73e-04 | 0.006 | 0.085 | 0.021 | 5.68e-05 | 6.22e-04 | 0.092 |
| 21 | PAFAH1B3 | 1007 | 0.0193 | 4.92 | 6.22e-04 | 0.006 | 0.035 | 0.020 | 0.083 | 0.207 | 0.052 |
| 22 | pQ-ATXN3 | 1007 | 0.0191 | 4.86 | 6.96e-04 | 0.006 | 0.039 | 0.021 | 0.060 | 0.167 | 0.068 |
| 23 | TREM1 | 1007 | 0.0181 | 4.60 | 0.001 | 0.009 | 0.064 | 0.019 | 9.02e-04 | 0.007 | 0.008 |
| 24 | NPTX2 | 1007 | 0.0180 | 4.59 | 0.001 | 0.009 | 0.081 | 0.020 | 6.36e-05 | 6.59e-04 | 0.056 |
| 25 | NCAM1 | 1007 | 0.0177 | 4.50 | 0.001 | 0.010 | 0.084 | 0.022 | 1.64e-04 | 0.001 | 0.077 |
| 26 | Aβ42 | 1007 | 0.0175 | 4.46 | 0.001 | 0.011 | -0.055 | 0.021 | 0.011 | 0.051 | -0.181 |
| 27 | VGF | 1007 | 0.0168 | 4.28 | 0.002 | 0.014 | -0.051 | 0.020 | 0.013 | 0.058 | -0.060 |
| 28 | PPY | 1007 | 0.0167 | 4.26 | 0.002 | 0.014 | 0.073 | 0.019 | 1.28e-04 | 0.001 | 0.099 |
| 29 | IL1B | 1007 | 0.0154 | 3.92 | 0.004 | 0.025 | 0.063 | 0.021 | 0.002 | 0.015 | 0.085 |
| 30 | GBA | 1007 | 0.0153 | 3.88 | 0.004 | 0.026 | 0.076 | 0.021 | 2.34e-04 | 0.002 | 0.098 |
| 31 | GLP1R | 1007 | 0.0152 | 3.85 | 0.004 | 0.026 | 0.073 | 0.020 | 3.01e-04 | 0.002 | 0.081 |
| 32 | APOA1 | 1007 | 0.0143 | 3.61 | 0.006 | 0.038 | 0.062 | 0.022 | 0.004 | 0.025 | 0.073 |
| 33 | MAG | 1007 | 0.0137 | 3.46 | 0.008 | 0.048 | 0.071 | 0.021 | 7.92e-04 | 0.006 | 0.097 |
| 34 | EFEMP1 | 1007 | 0.0136 | 3.45 | 0.008 | 0.048 | 0.044 | 0.023 | 0.052 | 0.152 | 0.039 |
| 35 | MME | 1007 | 0.0127 | 3.23 | 0.012 | 0.068 | -0.050 | 0.022 | 0.020 | 0.083 | -0.094 |
| 36 | CD33 | 1007 | 0.0124 | 3.14 | 0.014 | 0.076 | 0.071 | 0.021 | 8.42e-04 | 0.006 | 0.070 |
| 37 | pRAB10-T73 | 1007 | 0.0124 | 3.13 | 0.014 | 0.076 | 0.067 | 0.021 | 0.001 | 0.009 | 0.103 |
| 38 | S100A12 | 1007 | 0.0123 | 3.12 | 0.015 | 0.076 | 0.056 | 0.021 | 0.008 | 0.041 | 0.056 |
| 39 | NRGN | 1007 | 0.0117 | 2.95 | 0.019 | 0.097 | -0.026 | 0.021 | 0.213 | 0.401 | -0.042 |
| 40 | PRKN | 1007 | 0.0114 | 2.88 | 0.022 | 0.108 | -0.027 | 0.021 | 0.199 | 0.393 | -0.038 |
| 41 | PSEN1 | 1007 | 0.0113 | 2.86 | 0.022 | 0.108 | 0.043 | 0.021 | 0.035 | 0.124 | 0.061 |
| 42 | IGF1R | 1007 | 0.0111 | 2.81 | 0.025 | 0.115 | 0.063 | 0.021 | 0.003 | 0.017 | 0.030 |
| 43 | APOH | 1007 | 0.0110 | 2.77 | 0.026 | 0.119 | -0.058 | 0.021 | 0.006 | 0.033 | -0.129 |
| 44 | MDK | 1007 | 0.0108 | 2.74 | 0.028 | 0.122 | 0.023 | 0.019 | 0.237 | 0.428 | 0.040 |
| 45 | KLK6 | 1007 | 0.0108 | 2.73 | 0.028 | 0.122 | 0.066 | 0.021 | 0.002 | 0.011 | 0.057 |
| 46 | EIF4EBP1 | 1007 | 0.0108 | 2.72 | 0.029 | 0.122 | 0.037 | 0.020 | 0.062 | 0.171 | 0.048 |
| 47 | FOLR1 | 1007 | 0.0105 | 2.66 | 0.031 | 0.130 | 0.062 | 0.020 | 0.002 | 0.015 | 0.030 |
| 48 | NGF | 1007 | 0.0105 | 2.66 | 0.032 | 0.130 | 0.035 | 0.020 | 0.079 | 0.202 | -0.016 |
| 49 | IGFBP7 | 1007 | 0.0104 | 2.62 | 0.034 | 0.136 | 0.044 | 0.019 | 0.022 | 0.088 | -0.024 |
| 50 | GPNMB | 1007 | 0.0101 | 2.55 | 0.038 | 0.149 | 0.061 | 0.021 | 0.004 | 0.022 | 0.042 |
| 51 | NPTX1 | 1007 | 0.0100 | 2.52 | 0.040 | 0.154 | 0.058 | 0.021 | 0.005 | 0.029 | 0.086 |
| 52 | LRRK2 | 1007 | 0.0097 | 2.46 | 0.044 | 0.162 | 0.024 | 0.021 | 0.245 | 0.433 | 0.059 |
| 53 | NT-proBNP | 1007 | 0.0097 | 2.45 | 0.044 | 0.162 | 0.045 | 0.018 | 0.012 | 0.056 | 0.038 |
| 54 | CRP | 1007 | 0.0097 | 2.45 | 0.044 | 0.162 | -0.039 | 0.021 | 0.055 | 0.157 | -0.103 |
| 55 | ATXN3 | 1007 | 0.0096 | 2.43 | 0.046 | 0.164 | 0.023 | 0.020 | 0.246 | 0.433 | 0.030 |
| 56 | CCL11 | 1007 | 0.0095 | 2.41 | 0.048 | 0.168 | 0.051 | 0.019 | 0.009 | 0.043 | 0.014 |
| 57 | SFTPD | 1007 | 0.0094 | 2.37 | 0.051 | 0.176 | -0.043 | 0.021 | 0.035 | 0.124 | -0.100 |
| 58 | APOE | 1007 | 0.0091 | 2.30 | 0.057 | 0.193 | -0.023 | 0.021 | 0.281 | 0.473 | -0.044 |
| 59 | CSF2 | 1007 | 0.0091 | 2.29 | 0.058 | 0.193 | -0.026 | 0.021 | 0.212 | 0.401 | -0.045 |
| 60 | SAA1 | 1007 | 0.0089 | 2.26 | 0.061 | 0.201 | 0.020 | 0.021 | 0.348 | 0.534 | 0.006 |
| 61 | YWHAZ | 1007 | 0.0087 | 2.18 | 0.069 | 0.223 | 0.042 | 0.021 | 0.044 | 0.141 | 0.110 |
| 62 | SOD1 | 1007 | 0.0086 | 2.17 | 0.071 | 0.224 | -0.003 | 0.020 | 0.889 | 0.937 | -0.003 |
| 63 | FAM3B | 1007 | 0.0086 | 2.16 | 0.072 | 0.224 | -0.045 | 0.021 | 0.033 | 0.123 | -0.089 |
| 64 | HSPB1 | 1007 | 0.0085 | 2.14 | 0.074 | 0.224 | -0.015 | 0.021 | 0.467 | 0.624 | -0.011 |
| 65 | NEFH | 1007 | 0.0085 | 2.14 | 0.074 | 0.224 | 0.038 | 0.020 | 0.058 | 0.164 | 0.036 |
| 66 | SERPINA3 | 1007 | 0.0084 | 2.12 | 0.076 | 0.224 | 0.055 | 0.021 | 0.010 | 0.048 | 0.050 |
| 67 | CCL5 | 1007 | 0.0084 | 2.12 | 0.076 | 0.224 | -0.035 | 0.021 | 0.086 | 0.212 | -0.037 |
| 68 | POSTN | 1007 | 0.0083 | 2.10 | 0.079 | 0.229 | 0.054 | 0.022 | 0.015 | 0.064 | 0.023 |
| 69 | CHI3L1 | 1007 | 0.0082 | 2.08 | 0.081 | 0.232 | 0.039 | 0.019 | 0.037 | 0.128 | 0.002 |
| 70 | VCAM1 | 1007 | 0.0082 | 2.07 | 0.083 | 0.232 | 0.052 | 0.020 | 0.009 | 0.044 | 0.011 |
| 71 | RUVBL2 | 1007 | 0.0081 | 2.03 | 0.088 | 0.243 | 0.023 | 0.020 | 0.254 | 0.435 | 0.020 |
| 72 | SQSTM1 | 1007 | 0.0080 | 2.02 | 0.089 | 0.243 | -0.052 | 0.020 | 0.012 | 0.054 | -0.108 |
| 73 | EGFR | 1007 | 0.0078 | 1.96 | 0.099 | 0.267 | -0.046 | 0.021 | 0.032 | 0.120 | -0.111 |
| 74 | FGF2 | 1007 | 0.0076 | 1.91 | 0.106 | 0.278 | -0.013 | 0.021 | 0.531 | 0.684 | -0.013 |
| 75 | CSF1R | 1007 | 0.0075 | 1.90 | 0.108 | 0.278 | -0.037 | 0.021 | 0.082 | 0.206 | -0.116 |
| 76 | pLRRK2-S1292 | 1007 | 0.0075 | 1.90 | 0.108 | 0.278 | 0.046 | 0.021 | 0.025 | 0.099 | 0.089 |
| 77 | CST3 | 1007 | 0.0075 | 1.90 | 0.109 | 0.278 | 0.043 | 0.022 | 0.046 | 0.145 | -0.053 |
| 78 | CRH | 1007 | 0.0074 | 1.86 | 0.114 | 0.289 | 0.050 | 0.021 | 0.016 | 0.067 | 0.065 |
| 79 | TNFSF14 | 1007 | 0.0071 | 1.79 | 0.128 | 0.319 | -0.037 | 0.021 | 0.073 | 0.191 | -0.096 |
| 80 | PDLIM5 | 1007 | 0.0069 | 1.73 | 0.140 | 0.345 | -0.042 | 0.021 | 0.048 | 0.148 | -0.057 |
| 81 | DLG4 | 1007 | 0.0067 | 1.69 | 0.150 | 0.364 | 0.007 | 0.021 | 0.746 | 0.834 | 0.017 |
| 82 | MDH1 | 1007 | 0.0067 | 1.68 | 0.152 | 0.366 | 0.041 | 0.021 | 0.053 | 0.153 | 0.046 |
| 83 | NPTXR | 1007 | 0.0066 | 1.66 | 0.157 | 0.372 | -0.032 | 0.021 | 0.136 | 0.304 | -0.083 |
| 84 | NPY | 1007 | 0.0066 | 1.65 | 0.159 | 0.374 | 0.047 | 0.021 | 0.027 | 0.106 | 0.052 |
| 85 | IL18 | 1007 | 0.0064 | 1.62 | 0.167 | 0.383 | 0.039 | 0.020 | 0.051 | 0.152 | 0.033 |
| 86 | CXCL8 | 1007 | 0.0064 | 1.62 | 0.167 | 0.383 | -0.015 | 0.020 | 0.467 | 0.624 | -0.048 |
| 87 | PTPRS | 1007 | 0.0060 | 1.50 | 0.201 | 0.455 | 0.046 | 0.022 | 0.034 | 0.123 | 0.018 |
| 88 | LGALS3 | 1007 | 0.0058 | 1.46 | 0.211 | 0.467 | 0.035 | 0.022 | 0.107 | 0.251 | -0.009 |
| 89 | CD63 | 1007 | 0.0058 | 1.46 | 0.213 | 0.467 | -0.040 | 0.020 | 0.042 | 0.139 | -0.134 |
| 90 | DNM1L | 1007 | 0.0058 | 1.45 | 0.215 | 0.467 | -0.010 | 0.021 | 0.632 | 0.741 | -0.010 |
| 91 | CHIT1 | 1007 | 0.0058 | 1.45 | 0.216 | 0.467 | 0.010 | 0.021 | 0.621 | 0.738 | -0.012 |
| 92 | IL16 | 1007 | 0.0057 | 1.42 | 0.224 | 0.476 | 0.024 | 0.021 | 0.256 | 0.435 | 0.020 |
| 93 | pQ-HTT | 1007 | 0.0057 | 1.42 | 0.225 | 0.476 | 0.043 | 0.021 | 0.039 | 0.134 | 0.060 |
| 94 | HBA1 | 1007 | 0.0055 | 1.39 | 0.235 | 0.488 | 0.003 | 0.020 | 0.894 | 0.937 | -0.023 |
| 95 | UCHL1 | 1007 | 0.0055 | 1.39 | 0.235 | 0.488 | 0.028 | 0.021 | 0.185 | 0.377 | 0.048 |
| 96 | CCL3 | 1007 | 0.0054 | 1.37 | 0.244 | 0.500 | 0.015 | 0.020 | 0.461 | 0.624 | -0.053 |
| 97 | PGF | 1007 | 0.0054 | 1.35 | 0.249 | 0.503 | 0.035 | 0.021 | 0.093 | 0.223 | 0.024 |
| 98 | VEGFD | 1007 | 0.0053 | 1.34 | 0.254 | 0.503 | 0.044 | 0.022 | 0.044 | 0.141 | 0.103 |
| 99 | GOT1 | 1007 | 0.0053 | 1.34 | 0.255 | 0.503 | 0.036 | 0.021 | 0.087 | 0.213 | 0.017 |
| 100 | TNF | 1007 | 0.0053 | 1.33 | 0.258 | 0.503 | 0.022 | 0.021 | 0.292 | 0.476 | -0.051 |
| 101 | IL12p70 | 1007 | 0.0053 | 1.32 | 0.261 | 0.503 | 0.028 | 0.020 | 0.158 | 0.346 | 0.023 |
| 102 | PLAUR | 1007 | 0.0052 | 1.31 | 0.263 | 0.503 | 0.032 | 0.020 | 0.100 | 0.238 | -0.027 |
| 103 | MMP10 | 1007 | 0.0052 | 1.31 | 0.265 | 0.503 | 0.016 | 0.020 | 0.418 | 0.608 | -0.023 |
| 104 | CNTN2 | 1007 | 0.0052 | 1.31 | 0.265 | 0.503 | -0.019 | 0.021 | 0.355 | 0.534 | -0.064 |
| 105 | CXCL1 | 1007 | 0.0052 | 1.30 | 0.269 | 0.505 | -0.040 | 0.020 | 0.043 | 0.140 | -0.058 |
| 106 | S100B | 1007 | 0.0051 | 1.28 | 0.278 | 0.517 | 0.030 | 0.022 | 0.173 | 0.358 | 0.028 |
| 107 | FGF21 | 1007 | 0.0050 | 1.27 | 0.282 | 0.519 | 0.020 | 0.021 | 0.323 | 0.517 | 0.051 |
| 108 | IL17A | 1007 | 0.0050 | 1.26 | 0.286 | 0.521 | 0.041 | 0.021 | 0.052 | 0.152 | 0.050 |
| 109 | IL15 | 1007 | 0.0049 | 1.23 | 0.297 | 0.532 | 0.031 | 0.022 | 0.161 | 0.349 | 0.049 |
| 110 | CNDP1 | 1007 | 0.0049 | 1.22 | 0.299 | 0.532 | 0.040 | 0.023 | 0.072 | 0.191 | -0.044 |
| 111 | IL10 | 1007 | 0.0049 | 1.22 | 0.301 | 0.532 | 0.016 | 0.020 | 0.441 | 0.621 | 0.005 |
| 112 | pSNCA-129 | 1007 | 0.0048 | 1.22 | 0.303 | 0.532 | 0.031 | 0.020 | 0.127 | 0.291 | 0.057 |
| 113 | IL9 | 1007 | 0.0047 | 1.19 | 0.315 | 0.548 | 0.025 | 0.020 | 0.229 | 0.421 | 0.007 |
| 114 | L1CAM | 1007 | 0.0047 | 1.17 | 0.322 | 0.557 | -0.025 | 0.021 | 0.231 | 0.421 | -0.084 |
| 115 | SNCA | 1007 | 0.0046 | 1.15 | 0.330 | 0.566 | 0.028 | 0.020 | 0.164 | 0.351 | 0.045 |
| 116 | YWHAG | 1007 | 0.0045 | 1.14 | 0.337 | 0.572 | 0.039 | 0.022 | 0.079 | 0.202 | 0.113 |
| 117 | ARSA | 1007 | 0.0045 | 1.13 | 0.342 | 0.573 | 0.004 | 0.022 | 0.866 | 0.922 | 0.012 |
| 118 | FABP3 | 1007 | 0.0045 | 1.12 | 0.343 | 0.573 | 0.010 | 0.019 | 0.587 | 0.719 | -0.074 |
| 119 | CTNNB1 | 1007 | 0.0043 | 1.09 | 0.361 | 0.598 | 0.016 | 0.021 | 0.468 | 0.624 | 0.042 |
| 120 | REST | 1007 | 0.0043 | 1.08 | 0.367 | 0.602 | 0.017 | 0.022 | 0.422 | 0.608 | -0.018 |
| 121 | IFNG | 1007 | 0.0042 | 1.06 | 0.374 | 0.608 | 0.007 | 0.021 | 0.729 | 0.825 | -0.012 |
| 122 | EDA2R | 1007 | 0.0042 | 1.06 | 0.377 | 0.609 | -0.001 | 0.022 | 0.957 | 0.977 | -0.009 |
| 123 | PGK1 | 1007 | 0.0042 | 1.04 | 0.383 | 0.613 | 0.028 | 0.021 | 0.166 | 0.351 | 0.043 |
| 124 | F2R | 1007 | 0.0041 | 1.04 | 0.386 | 0.613 | -0.013 | 0.021 | 0.550 | 0.699 | -0.040 |
| 125 | HMOX1 | 1007 | 0.0041 | 1.02 | 0.395 | 0.622 | -0.003 | 0.021 | 0.878 | 0.930 | -0.043 |
| 126 | FCGR2A | 1007 | 0.0040 | 1.01 | 0.400 | 0.625 | -0.007 | 0.021 | 0.725 | 0.825 | -0.033 |
| 127 | IL6R | 1007 | 0.0040 | 1.00 | 0.406 | 0.625 | -0.027 | 0.021 | 0.204 | 0.396 | -0.085 |
| 128 | NFKB2 | 1007 | 0.0040 | 0.99 | 0.410 | 0.625 | -0.039 | 0.022 | 0.073 | 0.191 | -0.065 |
| 129 | KDR | 1007 | 0.0040 | 0.99 | 0.410 | 0.625 | -0.017 | 0.022 | 0.448 | 0.624 | -0.059 |
| 130 | Oligo-SNCA | 1007 | 0.0039 | 0.99 | 0.414 | 0.625 | 0.026 | 0.020 | 0.197 | 0.393 | 0.049 |
| 131 | Aβ40 | 1007 | 0.0039 | 0.98 | 0.415 | 0.625 | 0.028 | 0.021 | 0.198 | 0.393 | -0.007 |
| 132 | FLT1 | 1007 | 0.0039 | 0.97 | 0.423 | 0.631 | 0.002 | 0.021 | 0.943 | 0.967 | -0.031 |
| 133 | PARP1 | 1007 | 0.0037 | 0.93 | 0.444 | 0.658 | 0.025 | 0.022 | 0.244 | 0.433 | 0.036 |
| 134 | GDF15 | 1007 | 0.0037 | 0.92 | 0.453 | 0.666 | 0.016 | 0.017 | 0.344 | 0.534 | -0.060 |
| 135 | CCL2 | 1007 | 0.0036 | 0.90 | 0.463 | 0.676 | -0.020 | 0.020 | 0.320 | 0.516 | -0.053 |
| 136 | RAB29 | 1007 | 0.0035 | 0.88 | 0.473 | 0.683 | 0.012 | 0.021 | 0.578 | 0.719 | -0.007 |
| 137 | BCAN | 1007 | 0.0035 | 0.88 | 0.475 | 0.683 | 0.031 | 0.021 | 0.142 | 0.314 | 0.055 |
| 138 | ATXN2 | 1007 | 0.0035 | 0.87 | 0.478 | 0.683 | -0.008 | 0.021 | 0.695 | 0.796 | -0.002 |
| 139 | MSLN | 1007 | 0.0034 | 0.85 | 0.491 | 0.695 | 0.026 | 0.020 | 0.205 | 0.396 | 9.07e-04 |
| 140 | PRDX6 | 1007 | 0.0034 | 0.84 | 0.499 | 0.695 | 0.021 | 0.020 | 0.292 | 0.476 | 0.044 |
| 141 | IL7 | 1007 | 0.0033 | 0.84 | 0.501 | 0.695 | -0.011 | 0.021 | 0.587 | 0.719 | -0.005 |
| 142 | GDI1 | 1007 | 0.0033 | 0.83 | 0.505 | 0.695 | 0.021 | 0.021 | 0.334 | 0.527 | 0.042 |
| 143 | HPGDS | 1007 | 0.0033 | 0.83 | 0.509 | 0.695 | -0.015 | 0.020 | 0.457 | 0.624 | -0.034 |
| 144 | SMOC1 | 1007 | 0.0033 | 0.82 | 0.511 | 0.695 | 0.015 | 0.021 | 0.466 | 0.624 | 0.048 |
| 145 | HTT | 1007 | 0.0033 | 0.82 | 0.511 | 0.695 | -0.025 | 0.022 | 0.253 | 0.435 | -0.038 |
| 146 | PRDX5 | 1007 | 0.0029 | 0.74 | 0.566 | 0.756 | 0.016 | 0.021 | 0.423 | 0.608 | 0.034 |
| 147 | NTRK2 | 1007 | 0.0029 | 0.74 | 0.568 | 0.756 | 0.011 | 0.020 | 0.588 | 0.719 | 0.018 |
| 148 | C1q | 1007 | 0.0029 | 0.73 | 0.568 | 0.756 | 0.032 | 0.021 | 0.122 | 0.284 | 0.007 |
| 149 | IL6 | 1007 | 0.0029 | 0.72 | 0.577 | 0.758 | 0.025 | 0.020 | 0.207 | 0.396 | -0.011 |
| 150 | LDLR | 1007 | 0.0029 | 0.72 | 0.577 | 0.758 | -0.010 | 0.021 | 0.627 | 0.740 | -0.031 |
| 151 | CX3CL1 | 1007 | 0.0028 | 0.70 | 0.589 | 0.769 | 0.018 | 0.019 | 0.354 | 0.534 | -0.041 |
| 152 | PPBP | 1007 | 0.0027 | 0.69 | 0.602 | 0.780 | -0.020 | 0.021 | 0.348 | 0.534 | -0.011 |
| 153 | ENO2 | 1007 | 0.0027 | 0.67 | 0.611 | 0.786 | 0.029 | 0.021 | 0.174 | 0.358 | 0.053 |
| 154 | pTDP43-409 | 1007 | 0.0026 | 0.65 | 0.629 | 0.801 | 0.020 | 0.020 | 0.334 | 0.527 | 0.058 |
| 155 | TREM2 | 1007 | 0.0026 | 0.64 | 0.632 | 0.801 | 0.011 | 0.019 | 0.557 | 0.704 | -0.067 |
| 156 | ANXA5 | 1007 | 0.0026 | 0.64 | 0.634 | 0.801 | 0.031 | 0.021 | 0.133 | 0.302 | 0.038 |
| 157 | GRN | 1007 | 0.0025 | 0.63 | 0.642 | 0.805 | -0.005 | 0.021 | 0.813 | 0.885 | -0.042 |
| 158 | CST5 | 1007 | 0.0024 | 0.61 | 0.653 | 0.810 | 0.013 | 0.020 | 0.518 | 0.681 | -0.012 |
| 159 | CCL4 | 1007 | 0.0024 | 0.61 | 0.656 | 0.810 | 1.60e-04 | 0.020 | 0.994 | 0.997 | -0.020 |
| 160 | IL2 | 1007 | 0.0024 | 0.61 | 0.658 | 0.810 | -0.004 | 0.020 | 0.836 | 0.900 | -0.040 |
| 161 | DKK1 | 1007 | 0.0024 | 0.60 | 0.666 | 0.813 | -0.007 | 0.021 | 0.749 | 0.834 | -0.004 |
| 162 | PARK7 | 1007 | 0.0024 | 0.59 | 0.669 | 0.813 | -0.012 | 0.021 | 0.581 | 0.719 | 0.002 |
| 163 | AQP4 | 1007 | 0.0022 | 0.55 | 0.699 | 0.845 | 0.026 | 0.019 | 0.168 | 0.352 | 0.002 |
| 164 | NELL1 | 1007 | 0.0021 | 0.52 | 0.722 | 0.860 | 0.023 | 0.020 | 0.256 | 0.435 | 0.035 |
| 165 | CCL26 | 1007 | 0.0020 | 0.51 | 0.727 | 0.860 | -6.11e-04 | 0.020 | 0.976 | 0.986 | 0.005 |
| 166 | CCL17 | 1007 | 0.0020 | 0.51 | 0.727 | 0.860 | 0.019 | 0.021 | 0.355 | 0.534 | 0.008 |
| 167 | CCL13 | 1007 | 0.0020 | 0.51 | 0.729 | 0.860 | -0.025 | 0.021 | 0.222 | 0.413 | -0.062 |
| 168 | PDGFC | 1007 | 0.0020 | 0.50 | 0.739 | 0.866 | 0.006 | 0.022 | 0.786 | 0.867 | 0.009 |
| 169 | CD40LG | 1007 | 0.0019 | 0.49 | 0.746 | 0.870 | 0.005 | 0.021 | 0.823 | 0.891 | 0.015 |
| 170 | AGRN | 1007 | 0.0018 | 0.45 | 0.775 | 0.883 | -0.003 | 0.021 | 0.902 | 0.940 | -0.100 |
| 171 | GSDME | 1007 | 0.0018 | 0.44 | 0.779 | 0.883 | 0.015 | 0.020 | 0.464 | 0.624 | 0.010 |
| 172 | SFRP1 | 1007 | 0.0017 | 0.43 | 0.785 | 0.883 | -0.022 | 0.021 | 0.285 | 0.475 | -0.057 |
| 173 | DDC | 1007 | 0.0017 | 0.43 | 0.789 | 0.883 | -0.019 | 0.021 | 0.366 | 0.547 | -0.025 |
| 174 | CCL18 | 1007 | 0.0017 | 0.42 | 0.797 | 0.883 | -0.002 | 0.021 | 0.928 | 0.958 | -0.071 |
| 175 | GAS6 | 1007 | 0.0017 | 0.42 | 0.798 | 0.883 | 0.018 | 0.021 | 0.378 | 0.560 | -0.045 |
| 176 | TARDBP | 1007 | 0.0016 | 0.41 | 0.799 | 0.883 | 0.009 | 0.021 | 0.673 | 0.785 | 0.022 |
| 177 | FCN2 | 1007 | 0.0016 | 0.41 | 0.800 | 0.883 | -0.013 | 0.021 | 0.528 | 0.684 | -0.005 |
| 178 | AXL | 1007 | 0.0016 | 0.41 | 0.801 | 0.883 | 0.023 | 0.022 | 0.293 | 0.476 | -0.031 |
| 179 | CXCL13 | 1007 | 0.0016 | 0.41 | 0.802 | 0.883 | 0.005 | 0.020 | 0.788 | 0.867 | -0.009 |
| 180 | NLRP3 | 1007 | 0.0016 | 0.40 | 0.810 | 0.886 | 0.004 | 0.022 | 0.853 | 0.913 | 5.08e-04 |
| 181 | BDNF | 1007 | 0.0015 | 0.38 | 0.822 | 0.892 | 0.010 | 0.021 | 0.619 | 0.738 | 0.029 |
| 182 | PROS1 | 1007 | 0.0015 | 0.38 | 0.824 | 0.892 | 0.013 | 0.021 | 0.539 | 0.689 | -0.005 |
| 183 | NFKB1 | 1007 | 0.0014 | 0.34 | 0.851 | 0.916 | 0.005 | 0.021 | 0.809 | 0.885 | 0.011 |
| 184 | IL5 | 1007 | 0.0013 | 0.32 | 0.863 | 0.922 | -8.99e-04 | 0.020 | 0.963 | 0.978 | -0.071 |
| 185 | GDNF | 1007 | 0.0012 | 0.31 | 0.872 | 0.922 | -0.007 | 0.020 | 0.733 | 0.825 | -0.010 |
| 186 | PDGFRB | 1007 | 0.0012 | 0.31 | 0.875 | 0.922 | 0.013 | 0.021 | 0.527 | 0.684 | 0.014 |
| 187 | VEGFA | 1007 | 0.0012 | 0.30 | 0.880 | 0.922 | -0.002 | 0.022 | 0.929 | 0.958 | -0.030 |
| 188 | SLIT2 | 1007 | 0.0012 | 0.29 | 0.886 | 0.922 | 0.017 | 0.021 | 0.408 | 0.600 | -0.023 |
| 189 | CXCL10 | 1007 | 0.0011 | 0.28 | 0.889 | 0.922 | -0.016 | 0.020 | 0.431 | 0.611 | -0.053 |
| 190 | TIMP3 | 1007 | 0.0011 | 0.28 | 0.892 | 0.922 | -0.011 | 0.021 | 0.593 | 0.721 | -0.002 |
| 191 | IL13 | 1007 | 0.0011 | 0.28 | 0.894 | 0.922 | -6.88e-05 | 0.020 | 0.997 | 0.997 | -0.045 |
| 192 | CCL22 | 1007 | 0.0011 | 0.26 | 0.901 | 0.924 | -0.016 | 0.020 | 0.431 | 0.611 | -0.041 |
| 193 | RAB10 | 1007 | 0.0010 | 0.26 | 0.905 | 0.924 | 0.010 | 0.021 | 0.622 | 0.738 | 0.053 |
| 194 | STX4 | 1007 | 7.54e-04 | 0.19 | 0.944 | 0.959 | -0.015 | 0.021 | 0.482 | 0.638 | -0.010 |
| 195 | TEK | 1007 | 6.80e-04 | 0.17 | 0.954 | 0.963 | 0.012 | 0.022 | 0.597 | 0.722 | -0.029 |
| 196 | ITGAV | 1007 | 4.41e-04 | 0.11 | 0.979 | 0.982 | -0.008 | 0.020 | 0.684 | 0.788 | -0.027 |
| 197 | ICAM1 | 1007 | 4.05e-04 | 0.10 | 0.982 | 0.982 | 0.009 | 0.021 | 0.679 | 0.787 | -0.054 |

Supplementary Table S5. Adjacent-stage contrasts and longitudinal proteomic recapitulation

**A. Adjusted adjacent-stage contrasts among the 34 stage-associated non-tau proteins**

| **Protein** | **Domain** | **No plasma tau abnormality → Tau phosphorylation, Δ (q)** | **Tau phosphorylation → Early aggregation, Δ (q)** | **Early aggregation → Moderate aggregation, Δ (q)** | **Moderate aggregation → Advanced aggregation, Δ (q)** |
| --- | --- | --- | --- | --- | --- |
| GFAP | Inflammation | 0.88 (2.49e-38) | 0.26 (0.010) | 0.24 (0.015) | 0.48 (1.11e-10) |
| SPP1 | Inflammation | 0.29 (0.013) | 0.18 (0.215) | -0.10 (0.544) | 0.22 (0.095) |
| IL33 | Inflammation | 0.21 (0.067) | 0.08 (0.642) | 0.13 (0.457) | -0.01 (0.968) |
| MMP9 | Inflammation | 0.31 (0.012) | -0.12 (0.502) | 0.13 (0.468) | 0.03 (0.893) |
| MOG | Inflammation | 0.08 (0.571) | 0.17 (0.286) | 5.68e-5 (1.000) | 0.12 (0.397) |
| TREM1 | Inflammation | 0.24 (0.020) | 0.05 (0.801) | -0.03 (0.893) | -0.00 (0.992) |
| IL1B | Inflammation | 0.23 (0.067) | -0.17 (0.319) | 0.25 (0.145) | -0.06 (0.770) |
| NfL | Cytoskeletal abnormalities | 0.24 (0.013) | 0.28 (0.092) | -0.07 (0.770) | 0.54 (3.34e-10) |
| STMN2 | Cytoskeletal abnormalities | 0.10 (0.447) | 0.25 (0.104) | -0.03 (0.893) | 0.04 (0.821) |
| MAG | Cytoskeletal abnormalities | 0.02 (0.908) | 0.17 (0.275) | -0.05 (0.821) | 0.17 (0.238) |
| ACHE | Synaptic and neuronal network defects | 0.59 (4.16e-15) | 0.18 (0.263) | 0.26 (0.133) | 0.16 (0.271) |
| GAP43 | Synaptic and neuronal network defects | 0.30 (0.003) | 0.40 (0.002) | 0.00 (0.992) | 0.18 (0.159) |
| SNAP25 | Synaptic and neuronal network defects | 0.25 (0.013) | 0.23 (0.099) | -0.05 (0.821) | -0.00 (0.999) |
| BASP1 | Synaptic and neuronal network defects | 0.21 (0.095) | 0.11 (0.502) | 0.15 (0.332) | -0.15 (0.296) |
| APOA2 | Synaptic and neuronal network defects | -0.05 (0.757) | -0.24 (0.159) | 0.03 (0.893) | -0.13 (0.453) |
| NTRK3 | Synaptic and neuronal network defects | 0.19 (0.140) | -0.05 (0.821) | 0.18 (0.227) | 0.01 (0.983) |
| PAFAH1B3 | Synaptic and neuronal network defects | 0.29 (0.013) | -0.21 (0.159) | 0.27 (0.091) | -0.26 (0.067) |
| NPTX2 | Synaptic and neuronal network defects | 0.17 (0.159) | 0.05 (0.821) | 0.03 (0.893) | 0.10 (0.492) |
| NCAM1 | Synaptic and neuronal network defects | 0.16 (0.261) | 0.08 (0.612) | 0.07 (0.612) | 0.02 (0.908) |
| VGF | Synaptic and neuronal network defects | 0.09 (0.502) | -0.07 (0.751) | 0.03 (0.893) | -0.33 (0.010) |
| PPY | Synaptic and neuronal network defects | 0.11 (0.440) | 0.16 (0.446) | -0.13 (0.502) | 0.21 (0.104) |
| APOA1 | Synaptic and neuronal network defects | 0.12 (0.342) | 0.01 (0.945) | 0.21 (0.198) | -0.17 (0.261) |
| EFEMP1 | Synaptic and neuronal network defects | 0.22 (0.093) | 0.12 (0.447) | -0.05 (0.770) | -0.15 (0.261) |
| BACE1 | Pathological protein aggregation | 0.42 (4.34e-5) | -0.03 (0.882) | 0.27 (0.085) | 0.16 (0.172) |
| mHTT-exon1 | Pathological protein aggregation | 0.07 (0.620) | 0.23 (0.159) | 0.06 (0.801) | -0.05 (0.801) |
| pQ-ATXN3 | Pathological protein aggregation | 0.06 (0.770) | -0.31 (0.044) | 0.52 (7.57e-5) | -0.17 (0.227) |
| Aβ42 | Pathological protein aggregation | -0.33 (0.001) | 0.10 (0.492) | -0.04 (0.821) | 0.04 (0.861) |
| pRAB29-T71 | Aberrant proteostasis | 0.04 (0.821) | 0.27 (0.104) | 0.13 (0.550) | -0.09 (0.617) |
| pRAB12-S106 | Aberrant proteostasis | -0.02 (0.926) | 0.17 (0.342) | 0.14 (0.502) | 0.01 (0.945) |
| PSME1 | Aberrant proteostasis | 0.24 (0.061) | -0.11 (0.546) | 0.26 (0.106) | -0.19 (0.172) |
| GBA | Aberrant proteostasis | 0.07 (0.665) | -0.05 (0.821) | 0.25 (0.145) | 0.01 (0.945) |
| pPRKN-S65 | Altered energy homeostasis | 0.14 (0.215) | 0.10 (0.622) | 0.21 (0.275) | -0.05 (0.821) |
| GLP1R | Altered energy homeostasis | 0.19 (0.095) | 0.02 (0.926) | 0.01 (0.988) | 0.10 (0.502) |
| VSNL1 | Neuronal cell death | 0.21 (0.095) | 0.23 (0.067) | -0.10 (0.513) | 0.18 (0.159) |

Values are age- and sex-adjusted standardized differences between adjacent plasma stages; BH-FDR q values in parentheses were calculated across all 34×4=136 post-hoc contrasts. This analysis localises cross-sectional stage differences and should not be interpreted as within-person temporal transitions.

B. Longitudinal recapitulation among the 34 cross-sectionally stage-associated non-tau proteins

| **Protein** | **Predefined Neuro220 domain** | **Cross-sectional stage orientation** | **Annual slope, SD/year** | **SE** | **95% CI** | **Nominal P** | **BH-FDR q** |
| --- | --- | --- | --- | --- | --- | --- | --- |
| APOA2 | Synaptic and neuronal network defects | Decreasing | -0.247 | 0.031 | -0.307–-0.187 | 6.73e-16 | 2.29e-14 |
| ACHE | Synaptic and neuronal network defects | Increasing | 0.184 | 0.025 | 0.136–0.233 | 8.17e-14 | 1.39e-12 |
| NfL | Cytoskeletal abnormalities | Increasing | 0.112 | 0.018 | 0.076–0.147 | 6.72e-10 | 7.62e-09 |
| PPY | Synaptic and neuronal network defects | Increasing | 0.138 | 0.023 | 0.092–0.184 | 3.30e-09 | 2.81e-08 |
| PAFAH1B3 | Synaptic and neuronal network defects | Increasing | 0.117 | 0.030 | 0.059–0.176 | 8.43e-05 | 5.73e-04 |
| SPP1 | Inflammation | Increasing | 0.089 | 0.023 | 0.043–0.135 | 1.35e-04 | 7.67e-04 |
| TREM1 | Inflammation | Increasing | 0.067 | 0.018 | 0.031–0.103 | 2.59e-04 | 0.001 |
| IL33 | Inflammation | Increasing | 0.082 | 0.023 | 0.036–0.128 | 4.41e-04 | 0.002 |
| IL1B | Inflammation | Increasing | 0.117 | 0.035 | 0.049–0.185 | 7.73e-04 | 0.003 |
| GFAP | Inflammation | Increasing | 0.042 | 0.015 | 0.013–0.071 | 0.005 | 0.016 |
| Aβ42 | Pathological protein aggregation | Decreasing | 0.093 | 0.034 | 0.026–0.161 | 0.006 | 0.020 |
| BASP1 | Synaptic and neuronal network defects | Increasing | -0.072 | 0.027 | -0.126–-0.018 | 0.009 | 0.023 |
| GBA | Aberrant proteostasis | Increasing | 0.058 | 0.022 | 0.014–0.101 | 0.009 | 0.023 |
| MAG | Cytoskeletal abnormalities | Increasing | 0.048 | 0.018 | 0.012–0.084 | 0.010 | 0.023 |
| STMN2 | Cytoskeletal abnormalities | Increasing | 0.052 | 0.021 | 0.010–0.093 | 0.016 | 0.035 |
| GLP1R | Altered energy homeostasis | Increasing | 0.033 | 0.015 | 0.003–0.063 | 0.032 | 0.068 |
| EFEMP1 | Synaptic and neuronal network defects | Increasing | -0.087 | 0.043 | -0.170–-0.003 | 0.042 | 0.079 |
| SNAP25 | Synaptic and neuronal network defects | Increasing | -0.064 | 0.032 | -0.125–-0.002 | 0.043 | 0.079 |
| pQ-ATXN3 | Pathological protein aggregation | Increasing | 0.057 | 0.028 | 0.001–0.112 | 0.044 | 0.079 |
| VGF | Synaptic and neuronal network defects | Decreasing | 0.053 | 0.027 | -6.07e-04–0.106 | 0.053 | 0.090 |
| mHTT-exon1 | Pathological protein aggregation | Increasing | -0.056 | 0.034 | -0.122–0.010 | 0.098 | 0.159 |
| pRAB29-T71 | Aberrant proteostasis | Increasing | -0.040 | 0.025 | -0.089–0.009 | 0.110 | 0.170 |
| APOA1 | Synaptic and neuronal network defects | Increasing | -0.042 | 0.030 | -0.101–0.017 | 0.166 | 0.246 |
| NCAM1 | Synaptic and neuronal network defects | Increasing | -0.020 | 0.015 | -0.049–0.009 | 0.177 | 0.251 |
| GAP43 | Synaptic and neuronal network defects | Increasing | -0.027 | 0.027 | -0.079–0.025 | 0.310 | 0.379 |
| MMP9 | Inflammation | Increasing | 0.024 | 0.022 | -0.019–0.067 | 0.281 | 0.379 |
| MOG | Inflammation | Increasing | 0.029 | 0.028 | -0.027–0.084 | 0.312 | 0.379 |
| VSNL1 | Neuronal cell death | Increasing | 0.023 | 0.023 | -0.021–0.068 | 0.301 | 0.379 |
| NTRK3 | Synaptic and neuronal network defects | Increasing | 0.017 | 0.020 | -0.021–0.056 | 0.372 | 0.436 |
| pRAB12-S106 | Aberrant proteostasis | Increasing | -0.020 | 0.026 | -0.070–0.031 | 0.441 | 0.500 |
| BACE1 | Pathological protein aggregation | Increasing | -0.010 | 0.017 | -0.044–0.024 | 0.569 | 0.624 |
| pPRKN-S65 | Altered energy homeostasis | Increasing | -0.013 | 0.027 | -0.066–0.041 | 0.647 | 0.688 |
| NPTX2 | Synaptic and neuronal network defects | Increasing | -0.006 | 0.016 | -0.038–0.025 | 0.690 | 0.711 |
| PSME1 | Aberrant proteostasis | Increasing | 0.009 | 0.033 | -0.056–0.073 | 0.795 | 0.795 |

Among the 34 proteins, 15 changed longitudinally at BH-FDR q<0.05; 13 of these 15 changed in the direction consistent with the cross-sectional later-stage pattern (Supplementary Figure S3).

**C. Prespecified longitudinal stage-oriented domain scores**

| **Predefined domain** | **N proteins** | **Annual slope, SD/year** | **SE** | **95% CI** | **Nominal P** | **BH-FDR q** |
| --- | --- | --- | --- | --- | --- | --- |
| Synaptic and neuronal network defects | 13 | -0.007 | 0.009 | -0.024–0.011 | 0.454 | 0.567 |
| Inflammation | 7 | 0.064 | 0.012 | 0.040–0.087 | 1.32e-07 | 6.59e-07 |
| Aberrant proteostasis | 4 | -1.67e-04 | 0.017 | -0.033–0.033 | 0.992 | 0.992 |
| Pathological protein aggregation | 4 | 0.020 | 0.015 | -0.009–0.049 | 0.179 | 0.298 |
| Cytoskeletal abnormalities | 3 | 0.068 | 0.014 | 0.041–0.096 | 1.15e-06 | 2.87e-06 |

Primary domain-level inference was restricted to predefined Neuro220 domains containing ≥3 of the 34 proteins. Domain scores were equal-weight means of proteins standardized using the baseline stageable cohort; direction was frozen from the cross-sectional trajectory solution before longitudinal testing.

General abbreviations: Aβ, amyloid-β; AUC, area under the receiver-operating-characteristic curve; BD, brain-derived; BH-FDR, Benjamini-Hochberg false discovery rate; CDR-SB, Clinical Dementia Rating–Sum of Boxes; CI, confidence interval; eMTBR-tau243, endogenously cleaved microtubule-binding-region tau243; NPQ, NULISA Protein Quantification; PET, positron emission tomography; T1, amyloid-associated phosphorylated and secreted AD tau; T2, established AD tau proteinopathy.
